# A Genome-Wide Association Study in the Chilean admixed population reveals four new *loci* for COVID-19 severity and susceptibility

**DOI:** 10.64898/2026.10.13.26365424

**Authors:** Yolanda Espinosa-Parrilla, Macarena Fuentes-Guajardo, Laura Carvajal-Silva, Kevin Aguilar-Valdés, Iskra A. Signore, Tamara V. Arévalo, Daniela Zapata-Contreras, Gerardo Donoso, Paula Zuñiga-Pacheco, Héctor Valenzuela-Jorquera, Stefania A. Rosso Villanelo, Eduardo A. Tobar-Calfucoy, Cristian E. Yáñez, Francisca Carvajal, Juan M. Saez Hidalgo, Sergio Sanhueza, Romina A. Quiroga, Matías F. Martínez, Leslie C. Cerpa, Nelson M. Varela, Claudio L. Cappelli, Claudia P. Campillay-Véliz, Braulio Valdebenito-Maturana, Jonathan García-Araya, Rodolfo A. Santander, Jenssy Opazo, Rocío Retamales-Ortega, Pamela Bocchieri, Teresa A. Alarcón, Anita Plaza, Mauricio Moraga, Christian A. Muñoz, Cesar A. Echeverria, Cristina Dorador, Virginia A. Monardes-Ramírez, Carolina A. Selman, Estefanía Nova-Lamperti, Carola Otth, Benjamin M. Neale, Andrea Ganna, Luis A. Quiñones, María A. Lavanderos, Andrea X. Silva, Alicia Colombo, Ricardo A. Verdugo

**Affiliations:** School of Medicine, Magallanes University, Punta Arenas, Chile; Evolutionary and Medical Genomics of Magallanes (GEMMa), Center for Education, Healthcare and Investigation (CADI-UMAG), Magallanes University, Punta Arenas, Chile; Interuniversity Center on Healthy Aging (CIES), Punta Arenas, Chile; Chilean Hereditary Cancer Group (GCCH), Punta Arenas, Chile; Centre for Craniofacial and Regenerative Biology, Faculty of Dentistry Oral and Craniofacial Sciences, King’s College London, London, UK; Department of Basic and Clinical Oncology (DOBC), Faculty of Medicine, University of Chile, Santiago, Chile; Faculty of Medicine, Universidad de Atacama, Copiapó, Chile; Service of Anatomic Pathology, University of Chile Clinical Hospital HCUCH, Santiago, Chile; AUSTRAL-omics, Vice Rector’s Office for Research, Development and Artistic Creation, Austral University of Chile, Valdivia, Chile; Human Genetics Program, Institute of Biomedical Science (ICBM), Faculty of Medicine, University of Chile, Santiago, Chile; Department of Computer Science, Faculty of Physical and Mathematical Sciences, University of Chile, Santiago; Molecular and Translational Immunology Laboratory, Clinical Biochemistry and Immunology Department, Faculty of Pharmacy, University of Concepción, Concepción, Chile; Department of Pharmaceutical Sciences and Technology, Faculty of Chemical Sciences and Pharmacy, University of Chile, Santiago, Chile; Latin American Network for Implementation and Validation of Clinical Pharmacogenomics Guidelines (RELIVAF-CYTED), Santiago, Chile; Laboratory of Molecular Virology, Faculty of Marine Sciences and Biological Resources, University of Antofagasta, Antofagasta; Department of Epidemiology and Biostatistics, Computational Oncology Service, Memorial Sloan Kettering Cancer Center, New York, NY, United States; Laboratory of Microbial Complexity and Functional Ecology, Antofagasta Institute & Biotechnology Department, University of Antofagasta, Antofagasta; Emergency Public Assistance Hospital, Santiago, Chile; Emergency Medical Assistance Service (SAMU), Punta Arenas, Chile; Department of Anatomic Pathology, Faculty of Medicine, University of Chile, Santiago, Chile; Diagnostic Units, Arturo López Pérez Foundation, Santiago, Chile; Department of Medical Technology, Faculty of Health Sciences, University of Antofagasta, Antofagasta, Chile; ATACAMA OMICS, Laboratory of Molecular Biology and Genomics, Faculty of Medicine, University of Atacama, Copiapó, Chile; Clinical Laboratory of the Technical Area of Molecular Biology, Hospital del Salvador, Santiago; Institute of Clinical Microbiology, Faculty of Medicine, Austral University of Chile, Valdivia; Analytic and Translational Genetics Unit, Department of Medicine, Massachusetts General Hospital, Boston, MA, USA; Stanley Center for Psychiatric Research, Broad Institute of MIT and Harvard, Cambridge, MA, USA; Institute for Molecular Medicine Finland (FIMM), Helsinki Institute of Life Science (HiLIFE), University of Helsinki, Helsinki, Finland; Faculty of Medicine, University of Helsinki, Helsinki, Finland; Department of Basic Sciences, Faculty of Sciences, University of Bío-Bío, Chile

**Author notes:** These authors contributed equally to this work and share first authorship (Yolanda Espinosa-Parrilla, Macarena Fuentes-Guajardo). Passed away before article submission (Carola Otth).

**Keywords:** COVID-19, GWAS, Biorepository, Latin American populations, Underrepresented populations

## Abstract

**Background:** More than five years after the COVID-19 outbreak, the infection by the SARS-CoV-2 virus should still be considered as a global threat with highly heterogeneous clinical outcomes from asymptomatic to fatal depending on existing genetic and phenotypic inter-individual and inter-population variability. Our objective is to identify risk loci associated with COVID-19 in an admixed Latin American population.

**Methods:** We performed a genome-wide association study in the framework of the Chilean C19-GenoNet team, analyzing a cohort of 3,417 individuals recruited from the northernmost to the southernmost regions of the country, who presented variations in the severity of their COVID-19 clinical manifestations.

**Results:** Our investigation replicated the extensively reported association of several variants at the 3p21.31 *locus* with COVID-19 severity and found a suggestive association for a variant within the FOXP4 gene at 6p21.1 with COVID-19 hospitalization. Both associations appear to be shared with other populations. Importantly, our study identified three previously unreported loci associated with COVID-19 severity or hospitalization at 7q21.11, 16p13.12 and 6p25.3, and one additional *locus* at 4p13 associated with SARS-CoV-2 positivity, a proxy for infection susceptibility.

**Conclusions:** these new risk loci may represent population-specific effects in Chilean and, potentially, other Latin American admixed groups. Our study highlights the importance of expanding efforts to study underrepresented and genetically different populations, which may provide new clues on the genetic basis of human diseases and may help to anticipate global responses to emerging viral infections and future pandemics.

## BACKGROUND

On March 11, 2020, the World Health Organization declared the COVID-19 outbreak a pandemic and classified it a global health emergency until May 2023 (1,2). Although more than five years have passed since the onset of the pandemic, the SARS-CoV-2 infection should still be regarded as a global health threat (1). SARS-CoV-2 infection presents a wide spectrum of highly heterogeneous clinical outcomes, ranging from asymptomatic to fatal cases, reflecting genetic and phenotypic variability both among individuals and populations, that shapes the physiological host responses after viral infection (3). Genome-wide association studies (GWAS) have provided an opportunity to dissect the role of host genetics in the severity of COVID-19 manifestations and to identify genetic risk factors for susceptibility to infection by SARS-CoV-2, altogether leading to the identification of more than 50 *loci* associated with COVID-19 (4–8). Several of these *loci* correspond to key genes previously associated with immune response, viral pathogenesis, lung diseases and inflammatory conditions (7), with the 3p21.31 *locus* on chromosome 3 standing out as the first to be identified and the most significant and widely replicated (7,8). This *locus* encompasses several genes, including *LIMD1*, *SACM1L*, *SLC6A20*, *LZTFL1*, *CCR9*, *FYCO1*, *CXCR6*, *XCR1,* and *CCR3*, many of which may play a relevant role in the pathophysiology of COVID-19 (8). This is a region of around 50 kilobases that was inherited from Neanderthals and is carried by approximately 30% of people in South Asia, 8% of people in Europe, and 4% of the admixed American population (9). To identify causal genes at this *locus,* integrated genome-scale CRISPR loss-of-function screens and expression quantitative trait *loci* (eQTLs) analyses across diverse cell types and tissues has been performed, identifying *SLC6A20* and *CXCR6* as modulators of COVID-19 risk (10). In this regard, *SLC6A20* interacts with *ACE2,* the receptor of the SARS-CoV-2 Spike protein and the key host gene for viral entry (11), while *CXCR6* controls the localization of resident memory T lymphocytes in the lung and maintains a pool of airway resident memory T cells, which are crucial for defense against respiratory pathogens (12). Additionally, it has been shown that the *LZTFL1* gene at the 3p21.31 *locus* is expressed in the lung epithelium regulating the epithelial-mesenchymal transition, a relevant infection-response pathway in patients with COVID-19. These findings suggest that *LZTFL1* could be a potential therapeutic target (13). Another early-identified *locus* associated with COVID-19 severity is the 9q34.2 *locus*, detected in a study including Spanish and Italian patients with respiratory failure (8). This region encompasses the *ABO* gene, suggesting the involvement of blood-group-related mechanisms in disease severity.

Nevertheless, the association at the 9q34 region with COVID-19 severity has not been shown to be as robust as the one at the 3p21.31 region, nor as widely replicated (4,14,15). Another highly consistent genetic association with COVID-19 severity is the *FOXP4* region (6,16). *FOXP4* is a transcriptional repressor that modulates the expression of lung-specific genes involved in epithelial injury response and mucus production (17). Allelic variants in this gene and in its neighbouring lncRNA gene *FOXP4-AS*, which upregulates *FOXP4*, have been identified as risk factors for COVID-19 hospitalization (6,16). Among the most significant, other widely reported associations include the 21q22.11 *locus* (rs17860169 in *IFNAR2*), previously associated with risk of hospitalization (6,7), the 9p13 *locus* (rs10813976 next to *AQP7*), functionally linked to aquaporin regulation, and the 19q13 *locus* (rs77127536 next to *UPK1A*), an intergenic eQTL in which *ARHGAP33*, encoding a Rho GTPase-activating protein, has been proposed as the most plausible candidate gene (6).

Despite the large amount of data generated since the emergence of COVID-19, most genetic studies are either exclusive for European and Asian populations (5,8,18,19) or are primarily composed of individuals from these two populations in trans-ancestry meta-analyses (14,16,20). Such approaches are valuable for finding common genetic risk variants that similarly affect multiple populations, but they may overlook variants with pronounced effects in specific, underrepresented groups (21). The largest of these studies was led by the COVID-19 Host Genetics Initiative (HGI) embracing 219,692 SARS-CoV-2-infected cases and over three million controls from populations with different ancestries. In its second update, the HGI study also included samples from the Chilean C19-GenoNet study reported here (7). Still, only two studies have investigated genetic risk factors for COVID-19 phenotypes using exclusively Latin American cohorts, and both were conducted in Brazilian populations (15,22). These two investigations replicated previously identified *loci*, but more interestingly, led to the identification of rare allelic variants in four COVID-19-related genes and a new COVID-19 risk locus within the *DSTYK* gene at the 1q32.1 region (15,22). In addition, one meta-analysis of 4,702 hospitalized cases of Latin American ancestry recruited by SCOURGE in five Latin America countries and Spain, identified two novel *loci* associated with COVID-19 hospitalization near the *BAZ2B* and *DDIAS* genes (4). Beyond the scarcity of studies, associating genetic variation with disease risk poses additional challenges in Latin Americans populations, since they cannot be considered a single ethnic group but rather a diverse set of related populations with different combinations of Iberian, Amerindian and African ancestry components (23). Chile is characterized by an admixed population, and no GWAS of COVID-19 phenotypes has been conducted exclusively in Chilean individuals. Furthermore, its ancestry varies remarkably across its geography, but its ancestral components reported are approximately 43% Amerindian, 55% European, and 2% sub-Saharan African (24).

Here we present the first GWAS findings from the Chilean C19-GenoNet study, the first of its class in the country in terms of geographic coverage and sample size, aiming to identify risk and severity *loci* associated with COVID-19 in the admixed Chilean population through a nation-wide collaboration network. The study included a cohort of 3,417 individuals from the northernmost to the southernmost regions of the country, with different levels of COVID-19 clinical severity. Our investigation confirmed the known association at 3p21.31 with COVID-19 severity and detected a suggestive signal at *FOXP4* related to COVID-19 hospitalization (Figure 2). Both results are shared with other populations (7). Importantly, our analyses also revealed three previously unreported loci: two at 16p13.12 and 7q21.11, associated with COVID-19 severity, and one at 6p25.3, associated with COVID-19 hospitalization. In addition, we detected one new locus, at 4p13, associated with SARS-CoV-2 positivity, a proxy of infection susceptibility. These new risk loci may be unique to Chilean and possibly other Latin American, admixed populations.

## MATERIAL AND METHODS

### Recruitment

The Chilean COVID-19 network of centers “C19-GenoNet” started in 2020 with the participation of 9 nodes distributed throughout Chile: University of Tarapacá, University of Antofagasta, University of Atacama, University of Chile, Salvador Hospital, Emergency Public Assistance Hospital, University of Concepción, Austral University of Chile and University of Magallanes (Figure 1, left panel). The recruitment, performed under the direct responsibility of the Biobank of Tissue and Fluids of the University of Chile (BTUCH) to ensure high standards of sample and data quality, started in September 2020 and concluded in May 2021, including a total of 2,129 participants. Details on the characteristics of the cohort and the biorepository data collection and processing are thoroughly reported elsewhere (25). Briefly, individuals aged 18 or older were enrolled in the study only if they provided written informed consent to participate and were either patients hospitalized for acute COVID-19 or subjects who attended one of the public campaigns carried out by each recruiting center (Figure 1, Workflow panel, section 1). Hospitalized patients had laboratory confirmation with test results available through electronic medical records. Non-hospitalized patients were required to have confirmation of SARS-CoV-2 infection, either by laboratory testing (SARS-CoV-2 qRT-PCR test, SARS-CoV-2 Rapid Antigen Test, Antibody Tests for COVID-19) or a clinical diagnosis (25). The diagnostic methods used in each cohort are summarized in Table S3. In addition, 572 patients from the Arturo López Pérez Foundation were included in the study. Furthermore, DNA samples and demographic data of 716 participants of the ChileGenomico Project were retrieved from the Biobank of the University of Chile to provide a representative population-based control cohort (26).

**Figure 1.**
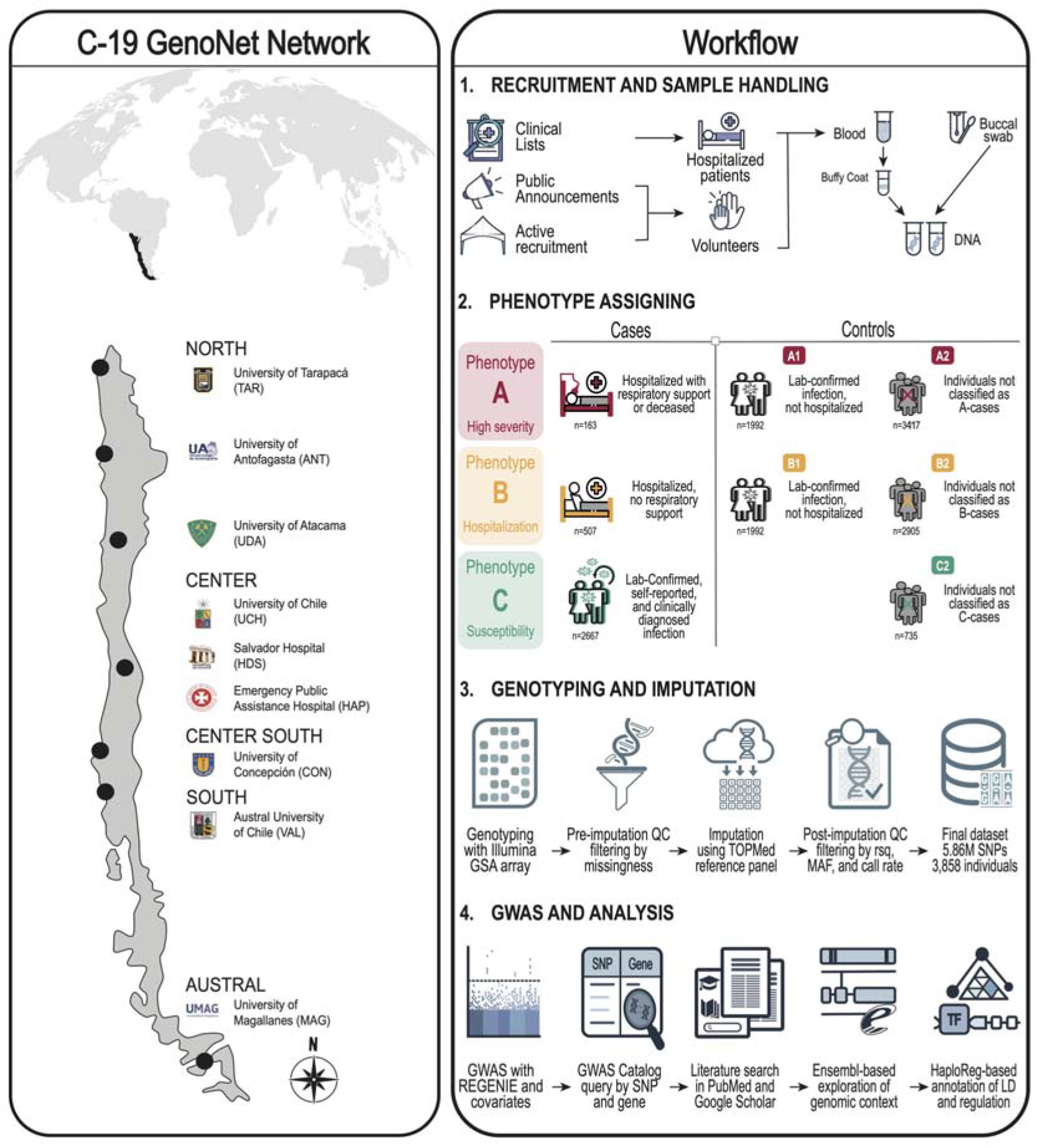
Graphic summary of the study. Left, map of Chile showing the major recruitment centers of the C19-GenoNet network. Right, workflow: 1, recruitment scheme and sample handling; 2, phenotype assignment, with the definition of cases and controls and the number of individuals per group; 3, genotyping and imputation workflow; 4, workflow for the GWAS and *in silico* annotation of candidate variants.

The project was approved by the Ethics Committee of the Faculty of Medicine of the University of Chile and was ratified by the local Ethics Committee at each participating site. The management of donor recruitment, samples, and data was carried out in compliance with national laws, international ethical-legal regulations, and good biobank practice (27–29).

### Sample collection and processing

For each participant, 6 mL of blood was collected in an EDTA tube, except for 55 cases, in which buccal swabbing was performed due to poor venous access. Blood samples were processed using a two-step process to obtain plasma (single spun) and buffy coat. DNA was purified from buffy-coat using either an EZNA blood DNA Mini Kit (Omega Bio-Tek) or QIAamp DNA Mini Blood (Qiagen, Inc.). Buccal swab samples were entirely used to purify DNA (Figure 1, Workflow panel, section 1).

### Assignment of Phenotypes

For the association analyses we assigned phenotypes as defined by the HGI (16), applying two COVID-19 severity phenotypes, A and B, and the C phenotype for partial susceptibility to SARS-CoV-2 infection (Figure 1, Workflow section 2). The A phenotypes represent the more severe outcomes. For the A1 and A2 categories, cases were defined as laboratory-confirmed SARS-CoV-2-infected individuals who were hospitalized for COVID-19 and received respiratory support or died (A Cases, n = 163). Additionally, we created two *wide* phenotypes, A3 and A4, with cases defined using the same criteria as in A1 and A2 but also including clinically diagnosed individuals (A-*wide* Cases, n = 336). Controls were different for each A phenotype as follows: for A1, controls were laboratory-confirmed SARS-CoV-2-infected individuals who were not hospitalized within 21 days after the test (A1 Controls, n = 1992). For A2, controls were all participants that were not classified as A cases (A2 Controls, n = 3249) (Figure 1, workflow section 2, first row). For A3, controls were laboratory-confirmed SARS-CoV-2-infected individuals who were hospitalized for COVID-19 but did not receive respiratory support and had not died (A3 Controls, n = 188). For A4, controls were laboratory-confirmed SARS-CoV-2-infected individuals who did not receive respiratory support and had not died, regardless of hospitalization (A4 Controls, n = 2238). The B phenotypes represent all hospitalized COVID-19 patients. For B1 and B2, cases were defined as laboratory-confirmed SARS-CoV-2-infected individuals who were hospitalized for COVID-19 regardless of respiratory support (B Cases, n = 507). Controls were different for each B phenotype: for B1, controls were the same as for A1 (B1 Controls, n = 1992), while for B2, controls were all participants who were not classified as B cases (B2 Controls, n = 2905) (Figure 1, workflow section 2, second row). Finally, the C phenotype represents SARS-CoV-2 partial-susceptibility. Cases were defined as laboratory confirmed SARS-CoV-2-infected individuals, clinically diagnosed or self-reported COVID-19 cases (C Cases, n = 2677). For C2, controls were all participants of population genomic survey in Chile that were recruited before the COVID-19 pandemic (C2 Controls, n = 735) (Figure 1, workflow section 2, third row).

### Genotyping, quality control, filtering and imputation

Genome-wide genotypes were generated by hybridization to Global Screening Arrays v3 (Illumina Inc.) in three batches processed in different core facilities. Only Single Nucleotide Polymorphisms (SNPs) were considered. Because the genotyping array captures primarily SNPs and only limited structural variation, indels were excluded at the genotyping stage and subsequently incorporated through imputation using the TOPMed reference panel. The raw genotypes underwent a first round of quality checks within each batch removing individuals with more than 2% and variants with more than 5% of missing data, respectively. SNPs with p-values <1×10^-06^ for Hardy-Weinberg equilibrium (HWE) and minor allele frequency (MAF) <0.01 were also removed. We standardized the assemblies in GRCh38 using the liftOver from the UCSC Genome Browser tool (30). The resulting datasets were merged with PLINK –merge and subsequently the same filters were applied on the merged dataset. A total of 444,740 variants and 3,417 individuals passed the quality tests. Subsequently, imputation of variants was performed using the “TOPMed Imputation Server” (version r2 online server) with a reference panel that included 97,256 individuals and 308,107,085 genetic variants distributed in the 22 autosomes and the X chromosome. A second round of quality checks were carried out on the imputed dataset using the PLINK/2.00 tool (31). These included variant filtering according to imputation quality score (rsq > 0.5), SNP call rate less than 95%, and sample call rate less than 98%. A total of 8,267,761 variants and 3,417 individuals passed the filters and quality controls (Figure 1, workflow section 3).

### Genome-Wide Association Study (GWAS)

GWAS were conducted to identify genetic variants associated with COVID-19 susceptibility and severity. Association analyses were performed using REGENIE v3.2.1 (32), a whole-genome regression method and modeled as: Phenotype ∼ age + age^2^ + sex + age*sex + 1-10 PCs + SNP, where PCs correspond to the principal components capturing population structure, and SNP represents the allele dosage of a SNP. Analyses were restricted to 5,863,724 autosomal variants with a minor allele frequency (MAF) ≥ 0.05 after standard quality control filtering and included 3,417 participants who had available data from at least one of the analyzed phenotypes. Instead of the widely used 5×10^-8^ threshold, genome-wide significance was set at 1.83×10^-7^, which is the empirical threshold proposed for Latin Americans based on 1000G data to account for their linkage disequilibrium (LD) structure (33). Associations with p-values below 1×10□□ that did not reach genome-wide significance were considered suggestive. Population structure correction was implemented by including the first ten principal components (PCs) derived from genotype data, reducing miscalculation due to ancestry differences. Manhattan and quantile-quantile (QQ) plots (Figure 2 and Figure S2) were generated using the R package qqman (34), and regional association plots were produced with LocusZoom v1.4 (35) to visualize local LD patterns around lead variants.

**Figure 2.**
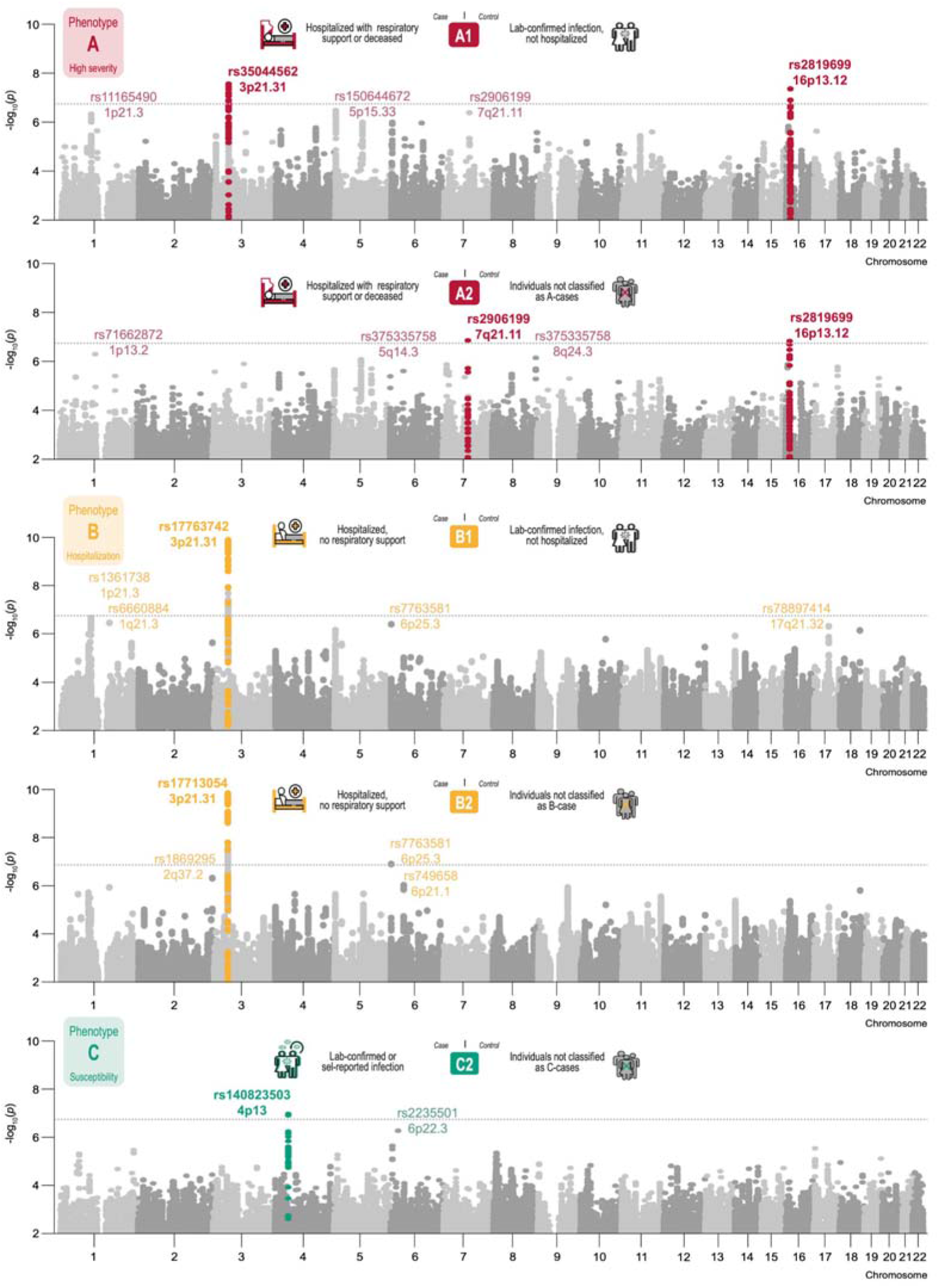
Manhattan plots of genome-wide association results for COVID-19 severity and susceptibility phenotypes in the Chilean admixed population. Results are shown for the A1, A2, B1, B2 and C2 phenotypes. Case and control definitions are summarized at the top of each panel. The dotted line represents the genome-wide significance threshold (1.83×10□□). Genome-wide significant loci are shown with coloured dots and labeled in bold with the rs identifier of the lead variant and the chromosomal region; the strongest suggestive associations are labeled in lighter colour.

### Linkage disequilibrium (LD) analysis

For the lead variant of each genome-wide significant locus, association results within a 1.4-Mb window (700 kb on each side) were extracted and pairwise LD (D′) was visualized as triangular heatmaps with LDBlockShow v1.41 (36), together with LocusZoom regional association plots (Figure 3). In addition, LD in a 10-kb window around each lead variant was examined with Haploview (37) and compared with that of the Iberian population in Spain (IBS) from the 1000 Genomes Project, retrieved from Ensembl (Figure S4).

**Figure 3.**
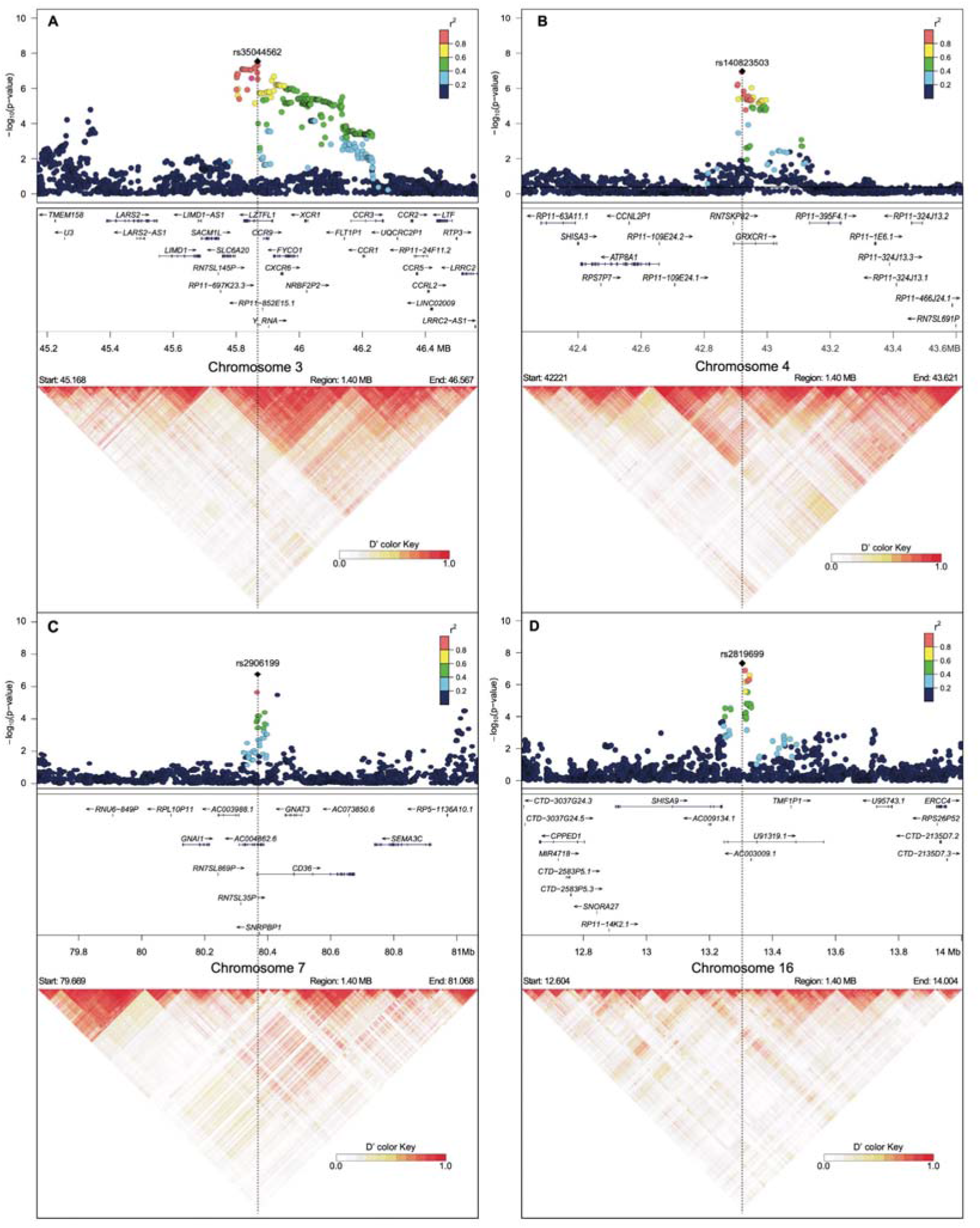
Regional association and linkage disequilibrium plots for genome-wide significant loci. (A) 3p21.31 (rs35044562, A1); (B) 4p13 (rs140823503, C2); (C) 7q21.11 (rs2906199, A2); (D) 16p13.12 (rs2819699, A1). Upper panels show LocusZoom regional association plots in a 1.4-Mb window centered on the lead variant (black diamond), with variants coloured by their r² with the lead variant and gene annotations below. Lower panels show LD heatmaps (D′) generated with LDBlockShow.

### Local Ancestry Inference (LAI) Analyses

LAI was carried out with Gnomix (38), using previously phased genotypes. The reference panel used was composed of 120 individuals from the 1000 Genomes project (40 European, 40 Asian, and 40 African) and 80 participants of the ChileGenomico project (40 Aymara and 40 Mapuche). After LAI, the per-individual information was merged with the respective phenotype information, allowing us to create the table that contains both the genetic and selected covariates information for statistical modelling using the same variables and model described earlier in the GWAS section and the functions “glm” and “anova” of the R statistical computing environment were used.

### Overview of candidate variants

We performed an *in silico* functional analysis using a structured multi-step approach to explore the potential functional implications of the GWAS signals. The SNPs associated with different acute COVID-19 phenotype groups were first checked using the GWAS Catalog, a curated database of published SNP-trait associations (39). The search was performed by SNP, gene and nearest genes within the genomic region showing association. These variants were then analyzed using HaploReg v4.2 (40) to investigate LD patterns, chromatin states, transcription factor binding motifs, and eQTL evidence across multiple tissues. Cis-eQTLs for the lead variants were retrieved from the GTEx Portal (release V10), where they are precomputed within ±1 Mb of the transcription start site of each gene (41). The prior associations found in the GWAS Catalog and the eQTL results are summarized in Table S1 and Figure S3. Further annotation of the SNPs was conducted using the Ensembl Genome Browser, which allowed us to map variants to nearby genes and assess their potential impact on gene structure and function, including coding and non-coding consequences. To visualize the genomic and epigenomic context of these variants, we employed the UCSC Genome Browser, integrating tracks such as gene models, regulatory elements, conservation scores, and ENCODE-derived chromatin data. Finally, PubMed was used to review existing literature related to the identified SNPs and candidate genes, supporting the interpretation of their possible biological roles in disease mechanisms. This integrative approach provided a comprehensive functional overview of candidate variants, helping to prioritize those with plausible regulatory or pathogenic significance for further experimental validation.

## RESULTS

The overall design of the Chilean C19-GenoNet study is based on the construction of a Chilean COVID-19 biorepository established by a network of nine nodes across five regions in Chile (Figure 1), from the northernmost (Arica y Parinacota) to the southernmost (Magallanes). The cohort included hospitalized COVID-19 patients as well as volunteer participants who attended public campaigns carried out by the recruiting centers. Individuals from ChileGenomico were also included as a population-based control group. Here, we present the first GWAS analysis performed after phenotyping and genotyping using 5,863,724 variants and the severity (A and B) and partial susceptibility (C) phenotypes defined by the COVID-19 HGI in a total of 3,417 Chilean individuals (Figure 1). Quantile-quantile plots for all phenotypes are shown in Figure S2. The strongest suggestive associations (p < 1×10□□) are labeled in Figure 2 and addressed in the Discussion.

### GWAS for COVID-19 high severity (A phenotypes)

Regarding phenotype A1 (Figure 1), where cases were confirmed SARS-CoV-2-infected individuals with severe COVID-19 and controls were confirmed SARS-CoV-2-infected individuals who were not hospitalized, the GWAS revealed two genome-wide significant associations (p<1.83×10^-07^, Table 1, Figure 2). In accordance with previous studies, we found an extensively reported association within an intron of the *LZTFL1* gene (Figure 3) at the 3p21.31 *locus* (chr3: 45867532, A/G; rs35044562, OR=4.02). This variant has been previously associated with COVID-19 hospitalization and respiratory support in the GWAS Catalog and is a cis-eQTL for several genes of the region, including *CXCR6*, *SLC6A20* and *CCR3* (Table S1, Figure S3). So far, this is the strongest genetic association with COVID-19 severity reported by multiple GWAS studies (7). In addition, we found a previously unreported association at the 16p13.12 *locus* (chr16:13304024; T/A; rs2819699, OR=2.28). The associated variant is located within an intron of the long non-coding RNA (lncRNA) gene *U91319.1* (Figure 3), which has been described as part of an immune-related lncRNA signature (42)*, a*nd next to the *SHISA9* gene, a regulator of short-term neuronal synaptic plasticity that has also been described as an autophagy cargo receptor (43).

**Table 1.**
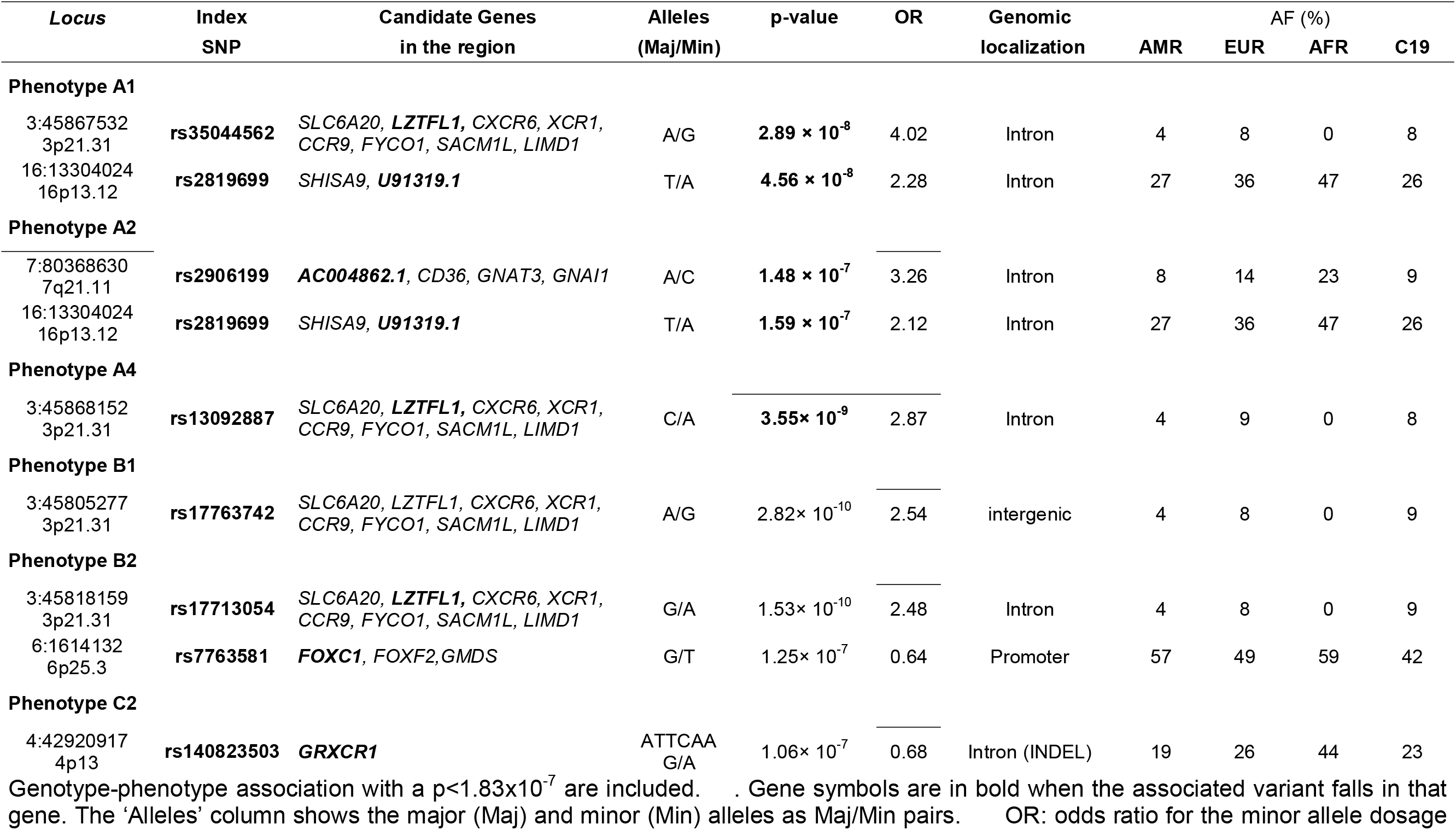
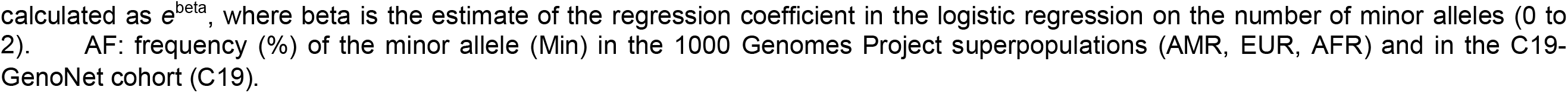
Results of the GWAS analysis in the Chilean C19-GenoNet study.

Regarding the A2 severity phenotype, cases followed the A1 definition, while controls were the individuals not classified as A cases. The analyses revealed two new genome-wide significant associations (Table 1, Figure 2), consisting of two associations that also arise for the A1 phenotype, the one at the 16p13.12 *locus* (rs2819699, OR=2.12) and the one at 7q21.11 (rs2906199, OR=3.26).

Regarding the A3 phenotype, cases were as A1 and A2 but also included clinical diagnosed COVID-19 individuals, and controls were hospitalized COVID-19 alive subjects that did not have respiratory support. Our analyses did not reveal genome-wide significant associations.

Finally, regarding the last severity A phenotype, the A4 phenotype, where cases were as A3 and controls were COVID-19 alive subjects that did not have respiratory support, the analyses revealed one genome-wide significant association (Table 1) (chr3: 45868152; C/A; rs13092887, OR=2.87) within the same *LZTFL1* gene at the 3p21.31 *locus* that rs35044562, the SNP associated with A1. A high LD (0.83) between both associated SNPs in *LZTFL1* was noted.

### GWAS for moderate severity and hospitalization (B phenotypes)

A hospitalization related phenotype, the B phenotype, where cases were COVID-19 patients that were hospitalized, regardless of the severity, was also analyzed. As for the B1 phenotype, where controls were non-hospitalized COVID-19 subjects, a genome-wide highly significant association (Table 1, Figure 3) of an intergenic variant next to the *SLC6A20, LZTFL1, CXCR6, XCR1, CCR9, FYCO1, SACM1L, LIMD1* genes at 3p21.31 *locus* was found (chr 3: 45805277; A/G; rs17763742, OR=2.54). This is the same genomic region that has been largely associated with COVID-19 and that we also found associated with the A1 and A4 phenotypes, and the associated SNPs are also in strong LD (0.84). Interestingly, *SLC6A20* has been associated with the susceptibility to SARS-CoV-2 infection, critical illness and respiratory symptoms in several populations (4,20,44–46).

Regarding the B2 phenotype, where controls were individuals that were not classified as B cases, we found two genome-wide significant associations (Table 1). The first one, at the 3p21.31 locus (Figure 3), corresponds to an intronic variant within the ENSG00000288720 gene (chr 3: 45818159, G/A, rs17713054, OR=2.48), a variant in an enhancer very close to the *LZTFL1* gene and that is in high LD (0.83) with the rs13092887 variant associated with A4. The second one is a previously unreported association at 6p25.3 (Figure 2) with a protective variant in the promoter region of *FOXC1* (chr 6:1614132, G/T, rs7763581, OR=0.64).

### GWAS for SARS-CoV-2 partial-susceptibility (C phenotype)

Finally, regarding the partial-susceptibility C2 phenotype, where cases were laboratory confirmed SARS-CoV-2 infected individuals, clinically confirmed COVID-19 or self-reported COVID-19 and controls were ChileGenomico pre-pandemic participants (Figure 1), a novel genome-wide significant association (Table 1, Figure 2) with a protective INDEL variant in the *GRXCR1* gene (Figure 3) at the 4p13 *locus* was found (chr 4: 42920917, ATTCAAG/A, rs140823503, OR=0.68), this *locus* in the vicinity of another *locus* (4p15.2) reported as associated with the presence of symptoms in a western Indian population (47), however, the associated variants in this population (rs12651262 and rs73246461) are not in LD with rs140823503.

### Local Ancestry Inference Analyses

After we selected the lead variants of the five genome-wide significant loci, we analyzed if there was a relationship between them and the Chilean local ancestry in these *loci*. We did not find statistically significant interactions of AMR ancestry for any of these loci (Table S2). However, in the case of rs35044562 at the 3p21.31 *locus*, although we did not observe individuals homozygous for the G risk allele, probably due to the low allele frequency, we observed a potentially higher risk for individuals carrying two Amerindian chromosomes and one risk allele (Figure S1), although this trend was not statistically significant (p=0.25). We did not find other potential interactions of AMR ancestry for any of the other genome-wide associated variants.

## DISCUSSION

Over the last years, GWAS have proved to be a useful tool for understanding the genetic basis of human susceptibility to infectious diseases by identifying *loci* associated with severity and progression of these diseases (48,49). Among these risk loci there are genes related to pathogen entrance, clearance, or persistence as well as genes related to the immune response of the host, such as those in the HLA genomic region (48). Here, we report an extensive national collaborative COVID-19 GWAS across different centers in Chile, the Chilean C19-GenoNet study, capturing the genetic diversity of the admixed Chilean population from the northernmost to the southernmost regions of the country. Our analyses revealed nine genome-wide significant associations with COVID-19-related phenotypes in five different *loci.* Only one of these *loci*, the one at 3p21.31, overlap with a previously and widely reported association, and the other four are new COVID-19 associated loci suggesting that they might harbour a population-specific effect. Three of them, the regions 7q21.11, 16p13.12 and 6p25.3 are associated with disease severity or hospitalization, while the locus at 4p13 is associated with SARS-CoV-2 positivity, a proxy of infection susceptibility. The association at 7q21.11 corresponds to a novel risk *loci* showing genome-wide significant association with A2 and a suggestive association with A1, remarkably, the associated variant is close to *CD36,* a transmembrane glycoprotein that is located on the cell surface of platelets and that may directly bind the envelope E protein of SARS-CoV-2, which can lead to increased platelet hyperactivity and the risk of the thrombotic events associated with COVID-19, this leads to propose CD36 as an actionable anti-thrombotic target for events related to SARS-CoV-2 infection (50).

Furthermore, mutations in the *CD36* gene have been found to be associated with an increased risk of cardiomyopathy, hyperlipidemia, and insulin resistance (10), conditions that can also be exacerbated by COVID-19. The analysis of LD blocks revealed a complex architecture for this region (Figure 3 and Figure S4), both facts suggesting a population-specific association at 7q21.11. Additionally, CD36 functions as both a signaling receptor and a fatty-acid transporter in multiple immune and non-immune cell types. Its dual roles help shape immune-cell function and fate, contributing to pathological processes such as atherosclerosis and tumor progression. These immunometabolic mechanisms may also modulate the host response to SARS-CoV-2 (51).

The association at 16p13.12 corresponds to a variant genome-wide significantly associated with both the A1 and A2 phenotypes, and it is in the intron of the lncRNA gene *U91319.1.* Very little is known about this gene except that it was described as part of a group of immune-related lncRNAs that serve as prognostic signature for cancer outcomes (42). It could be hypothesised a possible involvement of this lncRNA in the host immune response to the virus infection, a possibility that deserves further investigation. Noteworthy, the variant is close to *SHISA9*, a neuronal gene that has been recently described as an autophagy cargo receptor that mediates both antiviral and inflammatory responses in microglia and astrocytes at different stages during herpes simplex virus type 1 infection (43), whether this gene might have a similar role regarding other virus infections remains to be investigated. The third novel locus, at 6p25.3, corresponds to a protective variant (rs7763581) in the promoter region of *FOXC1* associated with COVID-19 hospitalization (B2 phenotype), which also showed a suggestive association with B1 (Figure 2). This variant has previously been associated with essential hypertension, high blood pressure and medication use for hypertension in the GWAS Catalog (Table S1), including in the multi-ancestry Million Veteran Program (52).

In the case of the fourth novel associated locus, it is at 4p13, corresponding to an indel variant that shows a protective effect, and lies within *GRXCR1*. This gene has previously been associated with deafness (53); however, no candidate genes related to virus infection susceptibility were identified in this genomic region. Either a pleiotropic effect of mutations in the *GRXCR1* gene or LD between the associated variant and the causative factor could underlie this association; the size of the LD block containing this SNP may make it challenging to identify the causative variant (Figure 3). Remarkably, the widely reported association at the 9q34 region with COVID-19 severity did not show up in our analysis, this could be indicating that the ABO *locus* is a population specific risk *locus*. Furthermore, the association of this *locus* with COVID-19 severity has been controversial and it appears to be related to SARS-CoV-2 infection susceptibility, with the ABO gene acting as virus receptor, rather than related to disease severity (15,54). Other previously and widely reported associations that could not be replicated with genome-wide significance in our cohort were these at 21q22.11, 9p13 and 19q13 *loci* (7). Likewise, specific variants in the *TMPRSS2*, a key gene that facilitates viral entry, have been reported as associated with COVID-19 susceptibility and severity across diverse populations worldwide, but were not identified here, suggesting that different host genetic backgrounds might contribute to differences in COVID-19 infection and severity (55,56). As far as we know, this is the first GWAS of COVID-19-related phenotypes conducted exclusively in the Chilean population and, besides the importance of replicating previously identified genomic associations, the discovery of potential novel risk *loci* that did not showed up in previous studies might be crucial in the follow-up efforts to identify population specific determinants of infectious diseases. This matches with the fact that Latin American populations are often underrepresented in genetic mapping studies (57), which missed the opportunities stemming from the high genetic diversity of these populations. Our results reinforce the idea that different host genetic backgrounds might contribute to differences in COVID-19 severity and susceptibility to infection. In fact, this genetic heterogeneity would be reflecting the complex of the host genetic architecture regarding different COVID-19 phenotypes with multiple genes involved in the response to the disease including a plethora of biological functions such as inflammatory pathways, immune cell activation and recruitment, pathogen recognition and entry and airway epithelia barriers, among others.

In addition to the genome-wide significant signals, several suggestive associations were observed in regions previously linked to COVID-19, supporting the consistency of our results with those reported in other populations. The most relevant is the intronic variant rs749658 within *FOXP4* at 6p21.1 (OR=1.50), suggestively associated with hospitalization (B2). This region was previously associated with COVID-19 severity in multi-ethnic but not in European-only meta-analyses (16), and *FOXP4*, involved in the response to injury of the airway epithelium (17), has also been associated with long COVID by the HGI (58). For the A4 phenotype, the variant rs607816 at 11q21, near *MAML2*, *FAM76B*, *MTMR2* and *CEP57* (OR=0.62), is in high LD (0.72) with rs590274, previously associated with severe COVID-19 (19); the splice variant rs3213689 within *ARHGAP44* at 17p12 (OR=2.19) lies near *ELAC2*, where rs12947402 has been associated with COVID-19 outcomes in a western Indian population (47), and near *MYOCD*, a gene associated with cardiac phenotypes; and the intronic variant rs75050680 within *GSG1L* at 16p12.1 (OR=2.14) is located in a gene where another SNP (rs755295), in low LD with ours (0.27), has been associated with the antibody response to SARS-CoV-2 vaccines (59). Regarding hospitalization (B1), the intergenic variant rs78897414 at 17q21.32 (OR=2.00) lies near *GOSR2*, which has been previously associated with hospitalization and cardiac COVID-19 phenotypes (58).

Other suggestive signals involve genes with plausible roles in the host response to SARS-CoV-2. For the A3 phenotype, the protective intronic variant rs28580753 (OR=0.27) within *AC007846.2* at 17p13.2 lies next to *NLRP1*, an inflammasome sensor that detects SARS-CoV-2 in lung epithelial cells through its cleavage by the viral 3CL protease (60). For the C2 phenotype, a protective missense variant in *GPLD1* at 6p22.3 (rs2235501, OR=0.50) was observed, and decreased expression of this gene has been correlated with COVID-19 severity (3). In addition, the variant rs2906199 near *CD36* at 7q21.11, genome-wide significant for A2, also showed a suggestive association with A1 (OR=3.27), as discussed above. The remaining suggestive associations, near *LINC02607* (1p21.3) and *IRX2* (5p15.33) for A1, at 1p13.2, *TMEM161B* (5q14.3) and *KCNK9* (8q24.3) for A2, at 6q21, 6q24.1, 8q11.22, 13q14.13 and 15q21.1 for A3, at 1p21.3 and 1q21.3 for B1, and near *AGAP1* (2q37.2) for B2, have not been previously related to COVID-19. Since none of these signals reached genome-wide significance and no replication cohort was available, they should be interpreted with caution and regarded as candidates for future studies.

One of the strongest points in our study comes to the fact that we constructed a Chilean COVID-19 biorepository based on biobanking standards, which allowed an effective collaborative national effort to recruit biospecimens and clinical data on COVID-19. This is the first Chilean initiative of this kind, and the cohort has already been part of HGI studies, being included in the GWAS analyses of COVID-19 and long covid (7,58). We expect that this biorepository will attract further international collaborations for the genetic and phenotypic studies on COVID-19 but also on the study of susceptibility to other infectious diseases. More than five years after the outbreak of COVID-19 there has been some lessons learned, as evidenced by the work done by the HGI consortia. The dissection of the host genetic factors associated with COVID-19 phenotypes was possible thanks to the joint effort of the community of human genetics researchers and their scientific collaboration by data sharing. This research model of international collaboration highlights what is possible for future genetic discoveries in emerging pandemics and/or forthcoming sanitary crises. In this sense, also national efforts, as the one reflected by our work, are of crucial relevance to finally dissect the underlying genetic basis of human disease considering interindividual and interpopulation heterogeneities.

A key important limitation of this study is the lack of a suitable replication sample for the novel COVID-19 associated *loci* and the reduced number of severe cases, which probably explains why some previously associated loci reached only suggestive significance, such as the *FOXP4* association with COVID-19 hospitalization (6,16), and why others, such as the loci near the *BAZ2B* and DDIAS genes reported in a meta-analysis of Latin American studies (4), were not replicated. Power estimates for the genome-wide significant associations, assuming a COVID-19 prevalence of 0.1, ranged from 0.39 to 0.99, with the lowest values for the A phenotypes (Table S4). Further efforts on the collection of more samples, particularly for local Chilean ancestral groups, such as the Mapuche and Aymara, as well as the finest dissection of the phenotypic data could provide new clues on the genetic basis of the host responses after SARS-CoV-2 infection from a wider population perspective, and, more outstandingly, it may help to anticipate the host responses to new emerging virus infections that may be the origin of future pandemics.

### Conclusions

The Chilean admixed population harbors genetic variation for COVID-19 susceptibility and severity. This study replicated the association between the *LZTFL1* polymorphism and severity and identified three novel loci at 7q21.11, 16p13.12 and 6p25.3 for severity or hospitalization. Additionally, a genetic association with SARS-CoV-2 positivity was identified at 4p13. Notably, the association at the ABO locus was not replicated in our study.

## Supporting information

Supplementary_material_GWAS_COVI19_Chile

## Data Availability

All data produced in the present study are available upon reasonable request to the authors

## ABBREVIATIONS

COVID-19: coronavirus disease 19
SARS-CoV-2: severe acute respiratory syndrome coronavirus-2 GWAS genome-wide association study
eQTLs: expression quantitative trait *loci*
HGI: host genetics initiative
SNPs: Single Nucleotide Polymorphisms
HWE: Hardy-Weinberg equilibrium
MAF: minor allele frequency
LD: linkage disequilibrium
PCs: principal components
QC: quality control
OR: odds ratio
LAI: local ancestry inference

## DECLARATIONS

### ETHICS APPROVAL AND CONSENT TO PARTICIPATE

The study was conducted under ethical and scientific standards and was approved by the relevant institutional ethical review boards. Informed consent was obtained from all individual participants included in the study. Further detailed information can be found in the text.

### CONSENT FOR PUBLICATION

Not applicable

### AVAILABILITY OF DATA AND MATERIALS

Data and biological samples may be made available upon request and after ethical and scientific evaluation and approval to ensure compliance with national laws and relevant policies of the biobanks, institutions and researchers involved in the biorepository. Individual-level ancestry estimates for European, African, Asian, and Amerindian origins are available along with demographic and clinically relevant variables. Inquiries should be directed to, who will provide a request form.

### AUTHORŚ CONTRIBUTIONS

The authors have contributed to this work as follows. Conceptualization: RAV; Methodology: JO, AP, RAQ, KAV; Investigation: YEP, KAV, TAA, LCC, MAL, NMV, CAE, AP, MFG, LGG, SGR, ELG, FC, JGA, JSH, JO, LCS, MFM, MM, CPCV, CAM, VMR, ENL, EAT, LAQ, SS, CAS, BVM, HVJ, DZC, PZP, PB, RAQ, RAS, AC, GD, IAS, AXS, TVA, RRO, SRV, CD, CEY, CLC, CO, BVM; Data curation: PB, GD, IAS, TVA, LCS, MFM, KAV; Formal analysis: LCS; Writing - Original draft: YEP, MFG, LCS; Writing – Review & Editing: RAV, LAQ, AC, IAS; Figures: IAS, LCS, BVM; Supervision: YEP, RAV, AC, AXS, MFG, ENL; Project administration: TVA; Funding acquisition: RAV, AC, YEP, MFM, LAQ, AXS, BMN, AG.

## ACKNOWLEDGEMENTS

Our deepest gratitude to all the participants of the C19-GenoNet Biorepository for their generous contribution to the science of Chile. We thank Eric S. Lander and Mark Daly for their significant support in securing the necessary funds for genotyping all DNA samples included in this study. We thank Annesi Giacaman, Claudio Verdugo, Cristóbal Verdugo, and Gerardo Acosta-Jamett for their assistance during recruitment. We also thank the technical and professional staff of the Biobank of Tissue and Fluids of the University of Chile for their commitment and support.

## REFERENCES

1. Wise J. Covid-19: WHO declares end of global health emergency. BMJ. 2023 May 9;381:1041.

2. Cucinotta D, Vanelli M. WHO Declares COVID-19 a Pandemic. Acta Biomed. 2020;91(1):157–60.

3. Overmyer KA, Shishkova E, Miller IJ, Balnis J, Bernstein MN, Peters-Clarke TM, et al. Large-Scale Multi-omic Analysis of COVID-19 Severity. Cell Syst. 2021 Jan 20;12(1):23–40.e7.

4. Diz-de Almeida S, Cruz R, Luchessi AD, Lorenzo-Salazar JM, de Heredia ML, Quintela I, et al. Novel risk loci for COVID-19 hospitalization among admixed American populations. Elife. 2024 Oct 3;13:RP93666.

5. Degenhardt F, Ellinghaus D, Juzenas S, Lerga-Jaso J, Wendorff M, Maya-Miles D, et al. Detailed stratified GWAS analysis for severe COVID-19 in four European populations. Hum Mol Genet. 2022 Nov 28;31(23):3945–66.

6. Ferreira LC, Gomes CEM, Rodrigues-Neto JF, Jeronimo SMB. Genome-wide association studies of COVID-19: Connecting the dots. Infect Genet Evol. 2022 Dec;106:105379.

7. COVID-19 Host Genetics Initiative. A second update on mapping the human genetic architecture of COVID-19. Nature. 2023 Sept;621(7977):E7–26.

8. Severe Covid-19 GWAS Group, Ellinghaus D, Degenhardt F, Bujanda L, Buti M, Albillos A, et al. Genomewide Association Study of Severe Covid-19 with Respiratory Failure. N Engl J Med. 2020 Oct 15;383(16):1522–34.

9. Zeberg H, Pääbo S. The major genetic risk factor for severe COVID-19 is inherited from Neanderthals. Nature. 2020 Nov;587(7835):610–2.

10. Kasela S, Daniloski Z, Bollepalli S, Jordan TX, tenOever BR, Sanjana NE, et al. Integrative approach identifies SLC6A20 and CXCR6 as putative causal genes for the COVID-19 GWAS signal in the 3p21.31 locus. Genome Biol. 2021 Aug 23;22(1):242.

11. Vuille-dit-Bille RN, Camargo SM, Emmenegger L, Sasse T, Kummer E, Jando J, et al. Human intestine luminal ACE2 and amino acid transporter expression increased by ACE-inhibitors. Amino Acids. 2015 Apr;47(4):693–705.

12. Wein AN, McMaster SR, Takamura S, Dunbar PR, Cartwright EK, Hayward SL, et al. CXCR6 regulates localization of tissue-resident memory CD8 T cells to the airways. J Exp Med. 2019 Dec 2;216(12):2748–62.

13. Downes DJ, Cross AR, Hua P, Roberts N, Schwessinger R, Cutler AJ, et al. Identification of LZTFL1 as a candidate effector gene at a COVID-19 risk locus. Nat Genet. 2021 Nov;53(11):1606–15.

14. Shelton JF, Shastri AJ, Ye C, Weldon CH, Filshtein-Sonmez T, Coker D, et al. Trans-ancestry analysis reveals genetic and nongenetic associations with COVID-19 susceptibility and severity. Nat Genet. 2021 June;53(6):801–8.

15. Pereira AC, Bes TM, Velho M, Marques E, Jannes CE, Valino KR, et al. Genetic risk factors and COVID-19 severity in Brazil: results from BRACOVID study. Hum Mol Genet. 2022 Sept 10;31(18):3021–31.

16. Niemi MEK, Karjalainen J, Liao RG, Neale BM, Daly M, Ganna A, et al. Mapping the human genetic architecture of COVID-19. Nature. 2021 Dec;600(7889):472–7.

17. Li S, Wang Y, Zhang Y, Lu MM, DeMayo FJ, Dekker JD, et al. Foxp1/4 control epithelial cell fate during lung development and regeneration through regulation of anterior gradient 2. Development. 2012 July 15;139(14):2500–9.

18. Esteban ME, Pino D, Romero-Lorca A, Novillo A, Gaibar M, Riancho JA, et al. Worldwide distribution of genetic factors related to severity of COVID-19 infection. Ann Hum Biol. 2024 Feb;51(1):2366248.

19. Słomian D, Szyda J, Dobosz P, Stojak J, Michalska-Foryszewska A, Sypniewski M, et al. Better safe than sorry—Whole-genome sequencing indicates that missense variants are significant in susceptibility to COVID-19. PLOS ONE. 2023 Jan 20;18(1):e0279356.

20. Pairo-Castineira E, Rawlik K, Bretherick AD, Qi T, Wu Y, Nassiri I, et al. GWAS and meta-analysis identifies 49 genetic variants underlying critical COVID-19. Nature. 2023;617(7962):764–8.

21. Li YR, Keating BJ. Trans-ethnic genome-wide association studies: advantages and challenges of mapping in diverse populations. Genome Medicine. 2014 Oct 31;6(10):91.

22. Secolin R, de Araujo TK, Gonsales MC, Rocha CS, Naslavsky M, Marco LD, et al. Genetic variability in COVID-19-related genes in the Brazilian population. Hum Genome Var. 2021;8:15.

23. Homburger JR, Moreno-Estrada A, Gignoux CR, Nelson D, Sanchez E, Ortiz-Tello P, et al. Genomic Insights into the Ancestry and Demographic History of South America. PLoS Genet. 2015 Dec;11(12):e1005602.

24. Eyheramendy S, Martinez FI, Manevy F, Vial C, Repetto GM. Genetic structure characterization of Chileans reflects historical immigration patterns. Nat Commun. 2015 Mar 17;6:6472.

25. Signore IA, Donoso G, Bocchieri P, Tobar-Calfucoy EA, Yáñez CE, Carvajal-Silva L, et al. The Chilean COVID-19 Genomics Network Biorepository: A Resource for Multi-Omics Studies of COVID-19 and Long COVID in a Latin American Population. Genes (Basel). 2024 Oct 22;15(11):1352.

26. Verdugo RA, Di Genova A, Herrera L, Moraga M, Acuña M, Berríos S, et al. Development of a small panel of SNPs to infer ancestry in Chileans that distinguishes Aymara and Mapuche components. Biological Research. 2020 Apr 16;53(1):15.

27. Annexe 2. WMA Declaration of Taipei on ethical considerations regarding health databases and biobanks:Adopted by the 53rd WMA General Assembly, Washington, DC, USA, October 2002 and revised by the 67th WMA General Assembly, Taipei, Taiwan, October 2016. Journal international de bioéthique et d’éthique des sciences. 2017 Nov 6;28(3):113–7.

28. Council for International Organizations of Medical Sciences. International ethical guidelines for biomedical research involving human subjects. Bull Med Ethics. 2002 Oct;(182):17–23.

29. Campbell LD, Astrin JJ, DeSouza Y, Giri J, Patel AA, Rawley-Payne M, et al. The 2018 Revision of the ISBER Best Practices: Summary of Changes and the Editorial Team’s Development Process. Biopreserv Biobank. 2018 Feb;16(1):3–6.

30. Perez G, Barber GP, Benet-Pages A, Casper J, Clawson H, Diekhans M, et al. The UCSC Genome Browser database: 2025 update. Nucleic Acids Res. 2025 Jan 6;53(D1):D1243–9.

31. Purcell S, Neale B, Todd-Brown K, Thomas L, Ferreira MAR, Bender D, et al. PLINK: a tool set for whole-genome association and population-based linkage analyses. Am J Hum Genet. 2007 Sept;81(3):559–75.

32. Mbatchou J, Barnard L, Backman J, Marcketta A, Kosmicki JA, Ziyatdinov A, Benner C, O’Dushlaine C, Barber M, Boutkov B, Habegger L, Ferreira M, Baras A, Reid J, Abecasis G, Maxwell E, Marchini J. Computationally efficient whole genome regression for quantitative and binary traits. Nat Genet. 2021 Jul;53(7):1097–103.

33. Kanai M, Tanaka T, Okada Y. Empirical estimation of genome-wide significance thresholds based on the 1000 Genomes Project data set. J Hum Genet. 2016 Oct;61(10):861–6.

34. Turner SD. qqman: an R package for visualizing GWAS results using Q-Q and manhattan plots. Journal of Open Source Software. 2018 May; 3(25), 731.

35. Pruim RJ, Welch RP, Sanna S, Teslovich TM, Chines PS, Gliedt TP, Boehnke M, Abecasis GR, Willer CJ. LocusZoom: regional visualization of genome-wide association scan results. Bioinformatics. 2010 Sep 15;26(18):2336–7.

36. Dong SS, He WM, Ji JJ, Zhang C, Guo Y, Yang TL. LDBlockShow: a fast and convenient tool for visualizing linkage disequilibrium and haplotype blocks based on variant call format files. Brief Bioinform. 2021 Jul 20;22(4):bbaa227.

37. Barrett JC, Fry B, Maller J, Daly MJ. Haploview: analysis and visualization of LD and haplotype maps. Bioinformatics. 2005 Jan 15;21(2):263–5.

38. Hilmarsson H, Kumar AS, Rastogi R, Bustamante CD, Mas Montserrat D, Ioannidis AG. High resolution ancestry deconvolution for next generation genomic data. bioRxiv. 2021. doi:10.1101/2021.09.19.460980.

39. Sollis E, Mosaku A, Abid A, Buniello A, Cerezo M, Gil L, et al. The NHGRI-EBI GWAS Catalog: knowledgebase and deposition resource. Nucleic Acids Res. 2023 Jan 6;51(D1):D977–85.

40. Ward LD, Kellis M. HaploReg v4: systematic mining of putative causal variants, cell types, regulators and target genes for human complex traits and disease. Nucleic Acids Res. 2016 Jan 4;44(D1):D877–81.

41. GTEx Consortium. The GTEx Consortium atlas of genetic regulatory effects across human tissues. Science. 2020 Sep 11;369(6509):1318–30.

42. Sun XX, Wen HQ, Zhan BX, Yang P. Identification of an Immune-Related LncRNA Prognostic Signature in Uterine Corpus Endometrial Carcinoma Patients. Clin Lab. 2021 Nov 1;67(11).

43. Zheng Y, Wang L, Liu Q, Xian H, Zhang C, Cai S, et al. Modulation of virus-induced neuroinflammation by the autophagy receptor SHISA9 in mice. Nat Microbiol. 2023 May;8(5):958–972.

44. Kousathanas A, Pairo-Castineira E, Rawlik K, Stuckey A, Odhams CA, Walker S, et al. Whole-genome sequencing reveals host factors underlying critical COVID-19. Nature. 2022 July;607(7917):97–103.

45. Alcalde-Herraiz M, Català M, Prats-Uribe A, Paredes R, Xie J, Prieto-Alhambra D. Genome-wide association studies of COVID-19 vaccine seroconversion and breakthrough outcomes in UK Biobank. Nat Commun. 2024 Oct 9;15(1):8739.

46. Kichaev G, Bhatia G, Loh PR, Gazal S, Burch K, Freund MK, et al. Leveraging Polygenic Functional Enrichment to Improve GWAS Power. Am J Hum Genet. 2019 Jan 3;104(1):65–75.

47. Pandit R, Singh I, Ansari A, Raval J, Patel Z, Dixit R, et al. First report on genome wide association study in western Indian population reveals host genetic factors for COVID-19 severity and outcome. Genomics. 2022 July;114(4):110399.

48. Tian C, Hromatka BS, Kiefer AK, Eriksson N, Noble SM, Tung JY, et al. Genome-wide association and HLA region fine-mapping studies identify susceptibility loci for multiple common infections. Nat Commun. 2017 Sept 19;8(1):599.

49. Malaria Genomic Epidemiology Network. Insights into malaria susceptibility using genome-wide data on 17,000 individuals from Africa, Asia and Oceania. Nat Commun. 2019 Dec 16;10(1):5732.

50. Tang Z, Xu Y, Tan Y, Shi H, Jin P, Li Y, et al. CD36 mediates SARS-CoV-2-envelope-protein-induced platelet activation and thrombosis. Nat Commun. 2023 Aug 21;14(1):5077.

51. Chen Y, Zhang J, Cui W, Silverstein RL. CD36, a signaling receptor and fatty acid transporter that regulates immune cell metabolism and fate. J Exp Med. 2022 Apr 19;219(6):e20211314.

52. Verma A, Huffman JE, Rodriguez A, Conery M, Liu M, Ho YL, et al. Diversity and scale: Genetic architecture of 2068 traits in the VA Million Veteran Program. Science. 2024 Jul 19;385(6706):eadj1182.

53. Schraders M, Lee K, Oostrik J, Huygen PLM, Ali G, Hoefsloot LH, et al. Homozygosity mapping reveals mutations of GRXCR1 as a cause of autosomal-recessive nonsyndromic hearing impairment. Am J Hum Genet. 2010 Feb 12;86(2):138–47.

54. D’Antonio M, Nguyen JP, Arthur TD, Matsui H, COVID-19 Host Genetics Initiative, D’Antonio-Chronowska A, et al. SARS-CoV-2 susceptibility and COVID-19 disease severity are associated with genetic variants affecting gene expression in a variety of tissues. Cell Rep. 2021 Nov 16;37(7):110020.

55. Cao Y, Li L, Feng Z, Wan S, Huang P, Sun X, et al. Comparative genetic analysis of the novel coronavirus (2019-nCoV/SARS-CoV-2) receptor ACE2 in different populations. Cell Discov. 2020;6:11.

56. Evans DG, van Veen EM, Byers H, Roberts E, Howell A, Howell SJ, et al. The importance of ethnicity: Are breast cancer polygenic risk scores ready for women who are not of White European origin? Int J Cancer. 2022 Jan 1;150(1):73–9.

57. Corpas M, Pius M, Poburennaya M, Guio H, Dwek M, Nagaraj S, et al.. Bridging genomics’ greatest challenge: The diversity gap. Cell Genom. 2025 Jan 8;5(1):100724.

58. Lammi V, Nakanishi T, Jones SE, Andrews SJ, Karjalainen J, Cortés B, et al. Genome-wide association study of long COVID. Nat Genet. 2025 June;57(6):1402–17.

59. Li P, Shi D, Shen W, Shi S, Guo X, Li J, et al. Pilot genome-wide association study of antibody response to inactivated SARS-CoV-2 vaccines. Front Immunol. 2022;13:1054147.

60. Planès R, Pinilla M, Santoni K, Hessel A, Passemar C, Lay K, et al. Human NLRP1 is a sensor of pathogenic coronavirus 3CL proteases in lung epithelial cells. Mol Cell. 2022 July 7;82(13):2385–2400.e9.

