## Supplementary_material_GWAS_COVI19_Chile for "A Genome-Wide Association Study in the Chilean admixed population reveals four new *loci* for COVID-19 severity and susceptibility"

Table S1. The prior association of each significant peak with COVID-19 or another trait found in the GWAS Catalog (https://www.ebi.ac.uk/gwas/) is shown. The main nearby genes for which the variants act as cis eQTLs are also shown (information taken from GTEx release V10: https://gtexportal.org/home/). GTEx cis-eQTLs are precomputed within a ±1 Mb window around the transcription start site of each gene.

| Phenotype | Chromosomal region | Index SNP | Associated phenotypes | Associated eQTL |
| --- | --- | --- | --- | --- |
| A1 | 3:45867532 | rs35044562 | - COVID-19 hospitalized vs population - COVID-19 respiratory support vs population | CXCR6, SLC6A20, CCR1, FLT1P1, CCR3, FYCO1, etc. |
| A1 /A2 | 16:13304024 | rs2819699 | Variant not found in Catalog | No eQTL found |
| A2 | 7:80368630 | rs2906199 | Variant not found in Catalog | ENSG00000232667 |
| A4 | 3:45868152 | rs13092887 | Variant not found in Catalog | No eQTL found |
| B1 | 3:45805277 | rs17763742 | Variant not found in Catalog | CXCR6, SLC6A20, CCR1, FLT1P1, CCR3, FYCO1 |
| B2 | 3:45818159 | rs17713054 | - COVID-19 severity, COVID-19 (critical illness vs population) - COVID-19 (hospitalized covid vs population) | CXCR6, SLC6A20, CCR1, FLT1P1, CCR3, FYCO1 |
| B2 | 6:1614132 | rs7763581 | - Essential hypertension - High blood pressure - Medication use for hypertension (number of purchases) | No eQTL found |

Table S2. Local ancestry at the lead variants of genome-wide significant loci. Counts of the effect allele (EAF) and of the non-effect allele (NEAF) are shown on chromosomes with Amerindian (AMR) and non-Amerindian (NAMR) local ancestry, together with the p-value of the comparison between ancestry backgrounds. EA, effect allele.

| Phenotype | Chromosomal region | Index SNP | EA | AMR EAF | NAMR EAF | AMR NEAF | NAMR NEAF | p-value |
| --- | --- | --- | --- | --- | --- | --- | --- | --- |
| A1 | 3:45867532 | rs35044562 | G | 911 | 1245 | 919 | 1237 | 0.83 |
| A1/A2 | 16:13304024 | rs2819699 | T | 1031 | 1125 | 987 | 1169 | 0.19 |
| A2 | 7:80368630 | rs2906199 | C | 1395 | 2022 | 1410 | 2007 | 0.73 |

Table S3. Diagnostic Methods of COVID-19 in participants.

| Diagnostic Method  Cohort | Laboratory confirmed | Clinical suspicion | Patient suspicion |
| --- | --- | --- | --- |
| Hospitalized | 243 |  |  |
| FALP | 565 |  |  |
| Outpatient | 1833 |  |  |

| 0, None | 1, Confirmed by a positive test (PCR or rapid test) | 2, Suspected by a health professional, not confirmed | 3, Self-suspected | NA |
| --- | --- | --- | --- | --- |
| 5 | 2636 | 42 | 2 | 732 |

Table S4. Statistical power for the genome-wide significant associations, estimated from the number of cases and controls, the allele frequency (AF, %) in the C19-GenoNet cohort, an assumed COVID-19 prevalence of 0.1 and the observed genotype relative risk. No genome-wide significant association was found for A3.

| Phenotype | Chromosomal region | Cases (n) | Controls (n) | AF | COVID19 prevalence | Genotype Relative Risk | Power |
| --- | --- | --- | --- | --- | --- | --- | --- |
| A1 | 3:45867532 | 163 | 1992 | 8.0 | 0.1 | 4.02 | 0.895 |
| A1 | 16:13304024 | 163 | 1992 | 26.7 | 0.1 | 2.28 | 0.537 |
| A2 | 7:80368630 | 163 | 3249 | 8.7 | 0.1 | 3.26 | 0.668 |
| A2 | 16:13304024 | 163 | 3249 | 26.2 | 0.1 | 2.12 | 0.385 |
| A3 | - | 336 | 188 |  |  |  |  |
| A4 | 3:45868152 | 336 | 2238 | 8.1 | 0.1 | 2.87 | 0.975 |
| B1 | 3:45805277 | 507 | 1992 | 8.6 | 0.1 | 2.54 | 0.995 |
| B2 | 3:45818159 | 507 | 2905 | 8.5 | 0.1 | 2.48 | 0.994 |
| B2 | 6:1614132 | 507 | 2905 | 41.9 | 0.1 | 1.56 | 0.758 |
| C | 4:42920917 | 2677 | 735 | 23.0 | 0.1 | 1.47 | 0.64 |

Figure S1. Interaction between the risk allele and Amerindian (AMR) local ancestry inferred by LAI for (A) rs35044562 (A1) and (B) rs140823503 (C2). The left panels show the distribution of cases (red) and controls (blue) per number of risk alleles, faceted by the number of local ancestry-inferred AMR chromosomes. The right panels show the average and error bars for the adjusted risk (RESID) by risk SNP genotype and by local AMR ancestry.

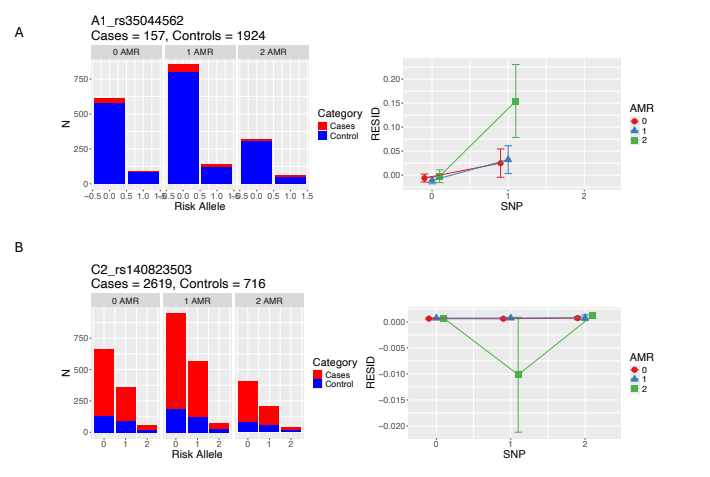

Figure S2. Q–Q plots for seven phenotypes estimated from GWAS analysis.

| phenotype A1  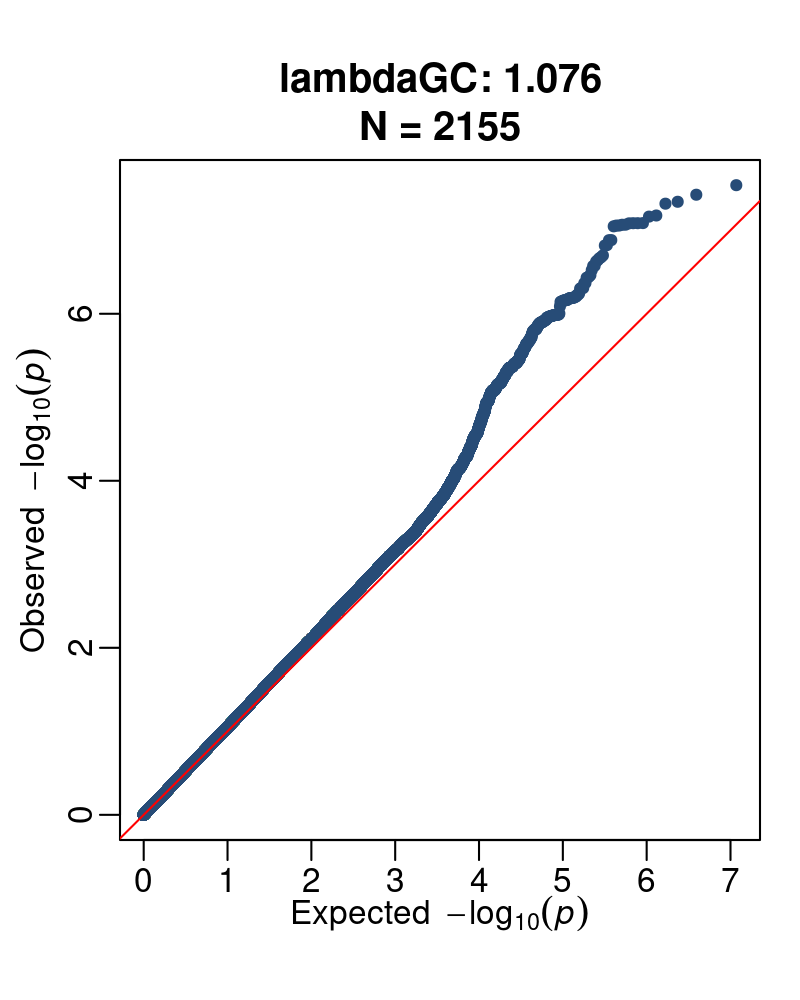 | phenotype A2  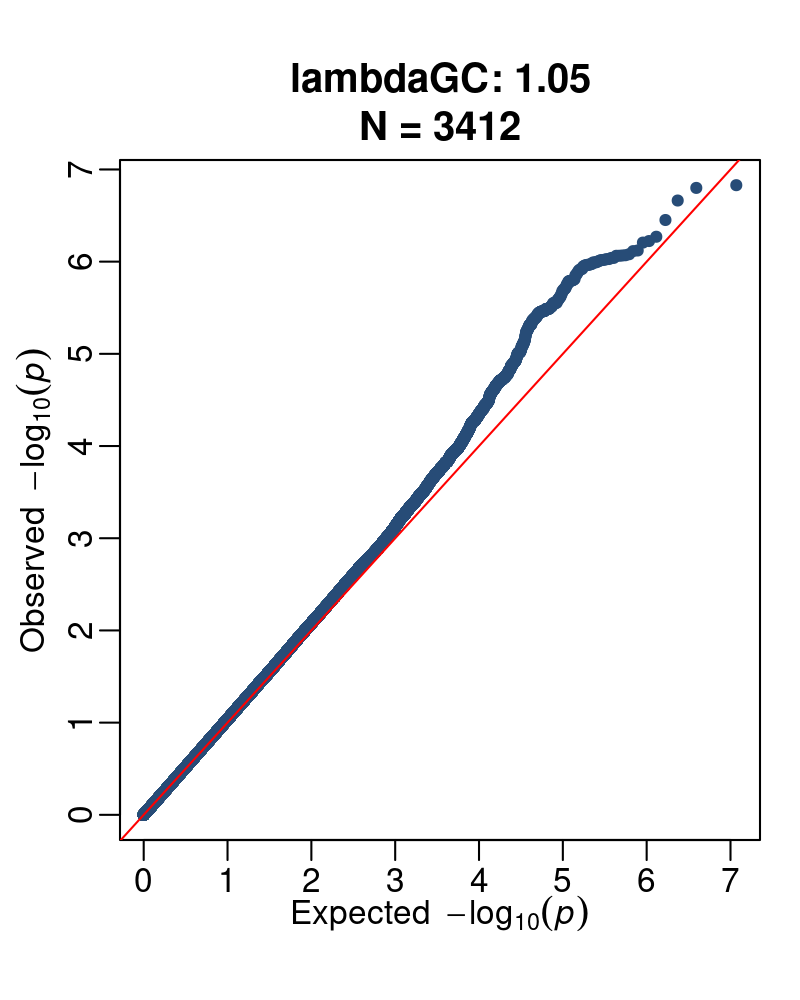 | phenotype A3  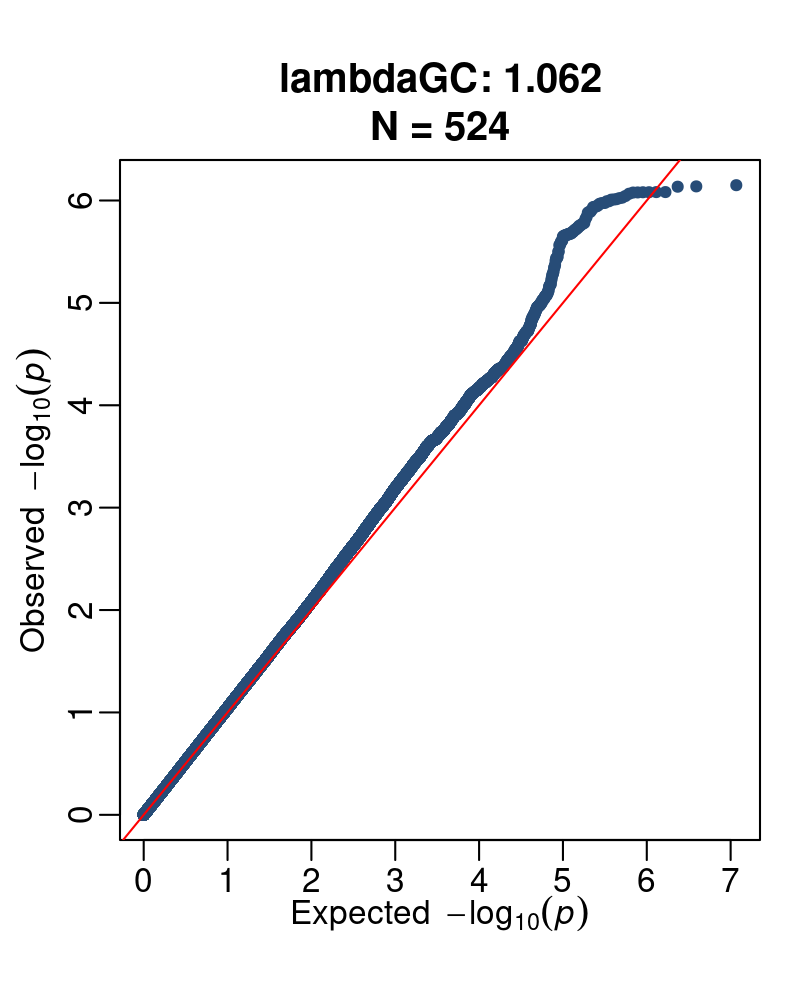 |
| --- | --- | --- |
| phenotype A4  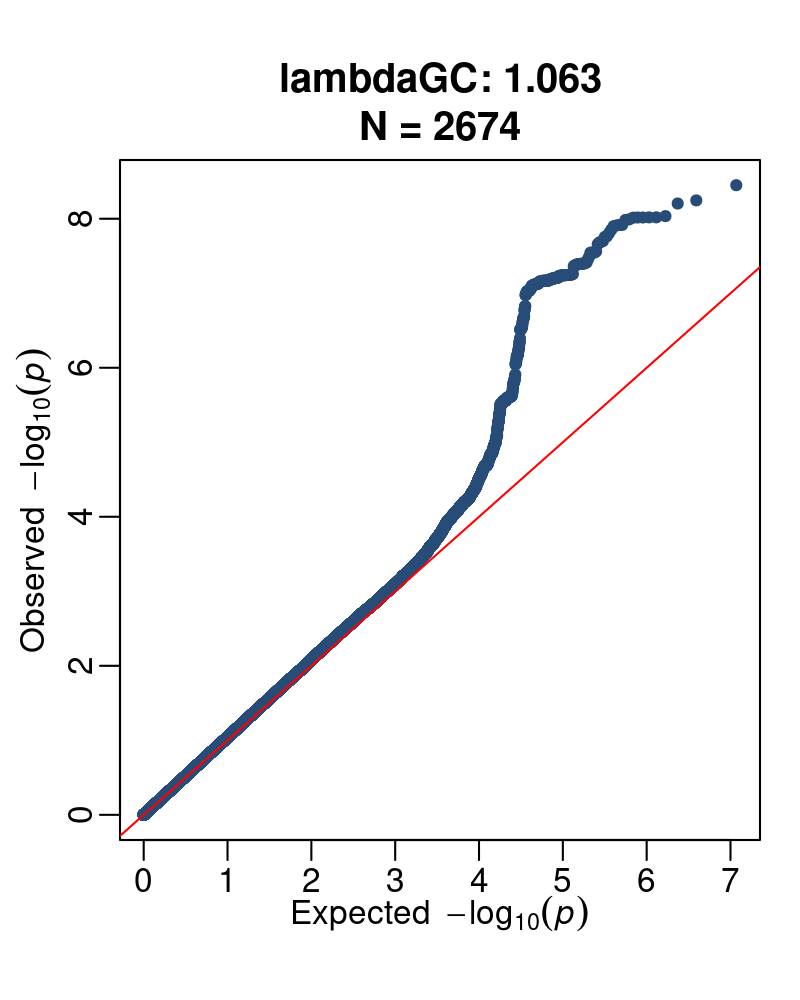 | phenotype B1  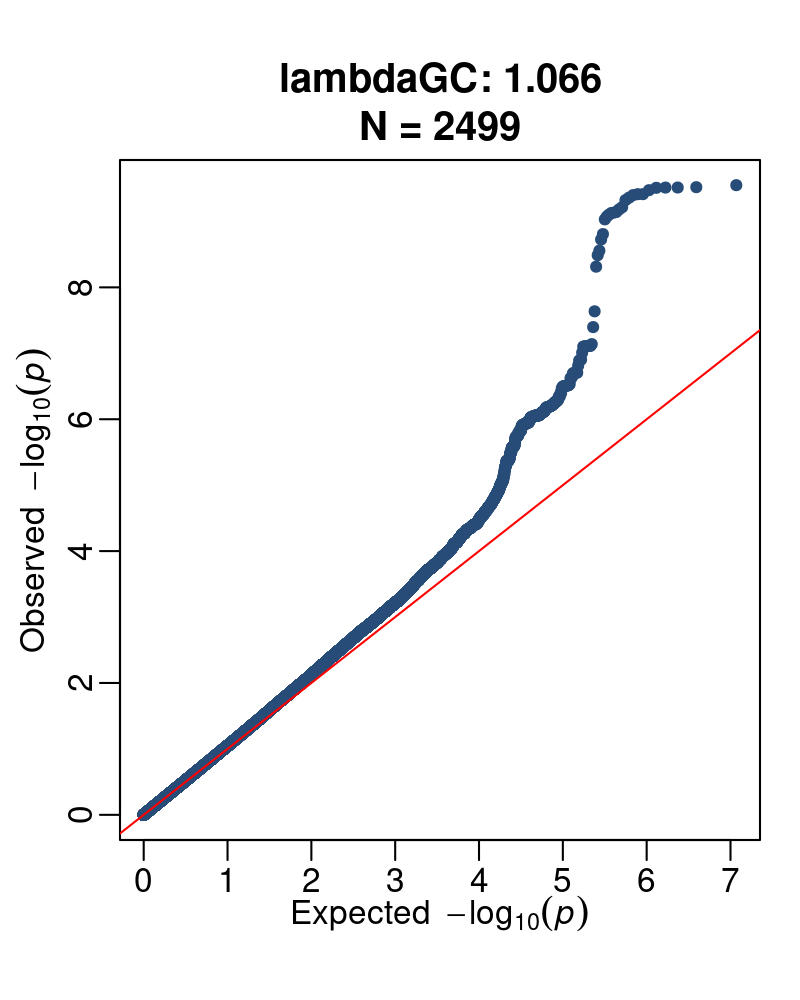 | phenotype B2  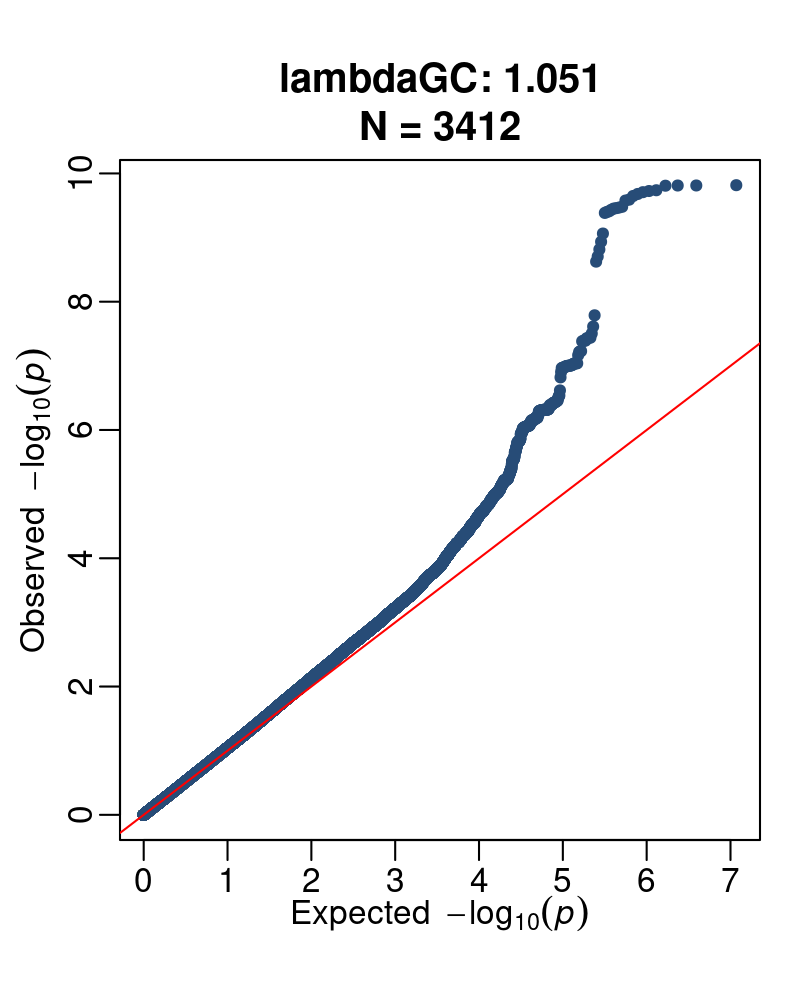 |
| phenotype C2  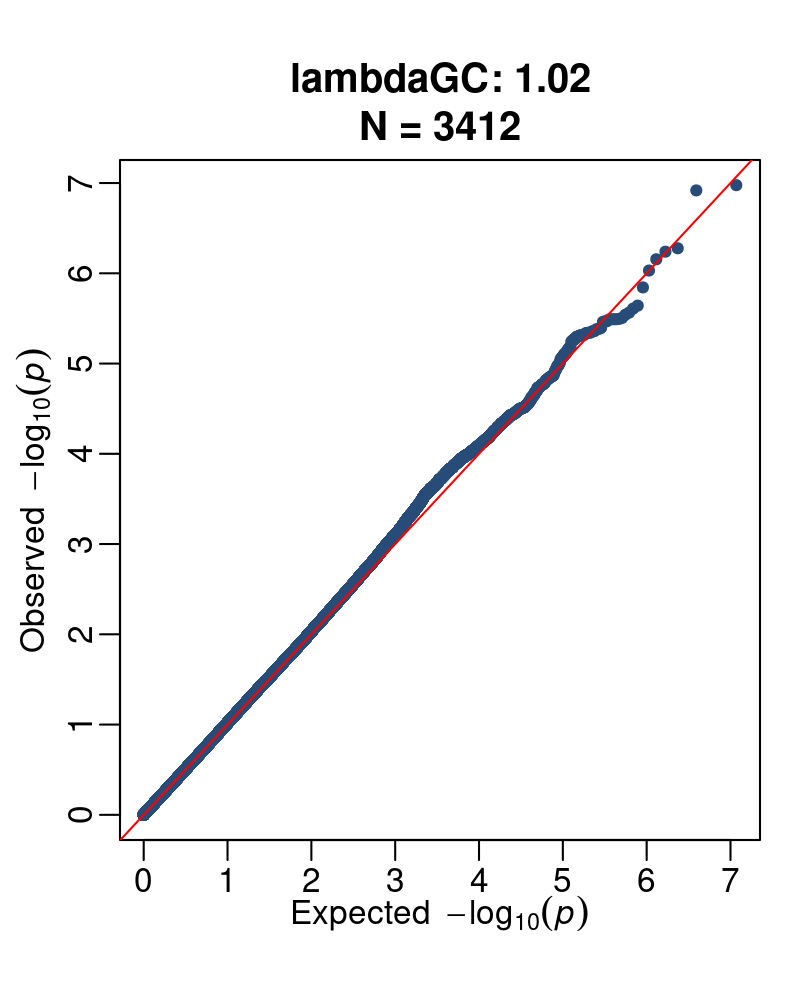 |  |  |

Figure S3. Main eQTL per peak. Single-tissue eQTL violin plots are shown for the significant variants. The tissue with the lowest p-value was selected to be displayed at each peak. (GTEx Analysis Release V10)

| 3:45867532 | | |
| --- | --- | --- |
| 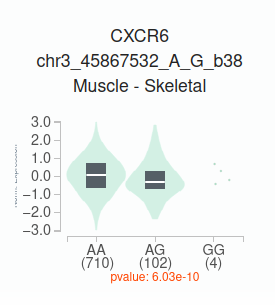 | 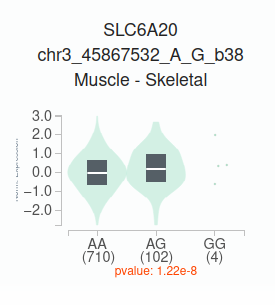 | 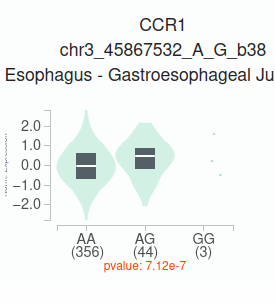 |
| 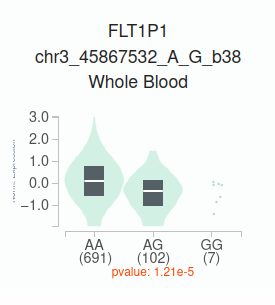 | 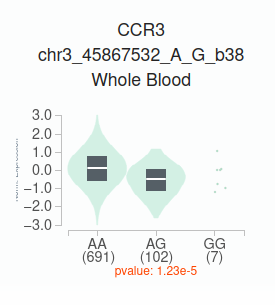 | 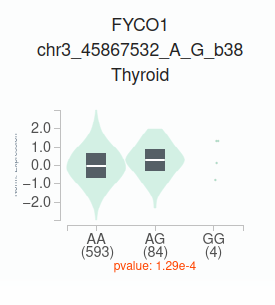 |
| 7:80368630 | | |
| 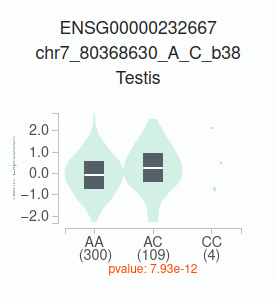 |  |  |
| 3:45805277 | | |
| 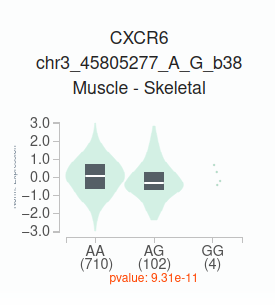 | 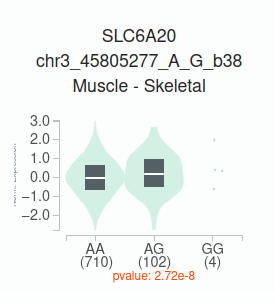 | 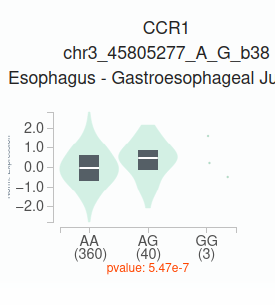 |
| 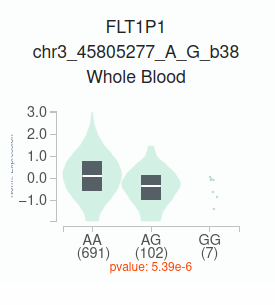 | 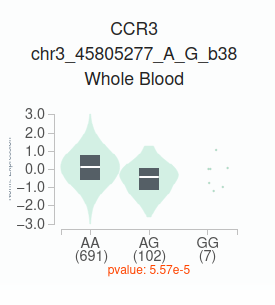 | 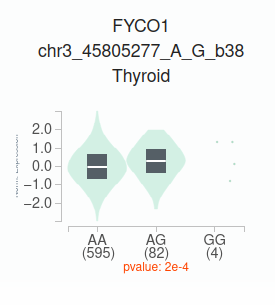 |
| 3:45818159 | | |
| 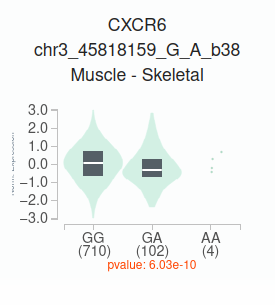 | 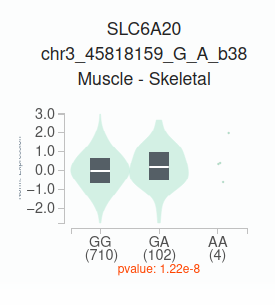 | 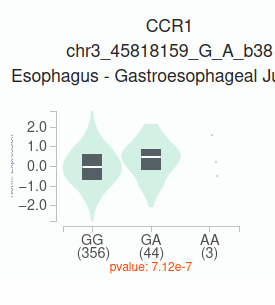 |
| 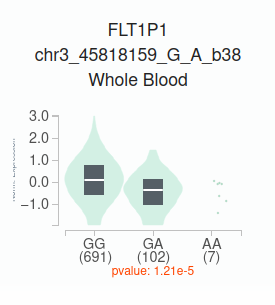 | 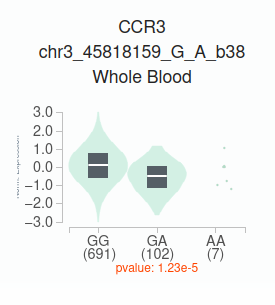 | 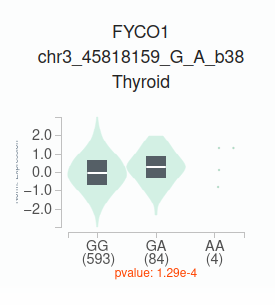 |

Figure S4. LD Plots from Haploview. Plots were generated for a 10,000bp window around the marker SNP of each peak. The windows compared the C19-GenoNet cohort with the Iberian population of Spain (taken from Ensembl).

| C19-GenoNet | Iberian (IBS, Spain) |
| --- | --- |
| A1 |  |
| 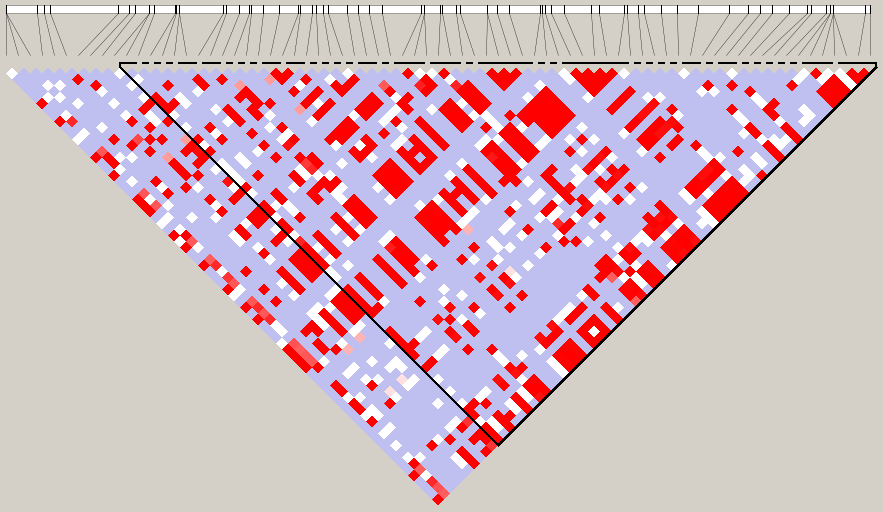 | 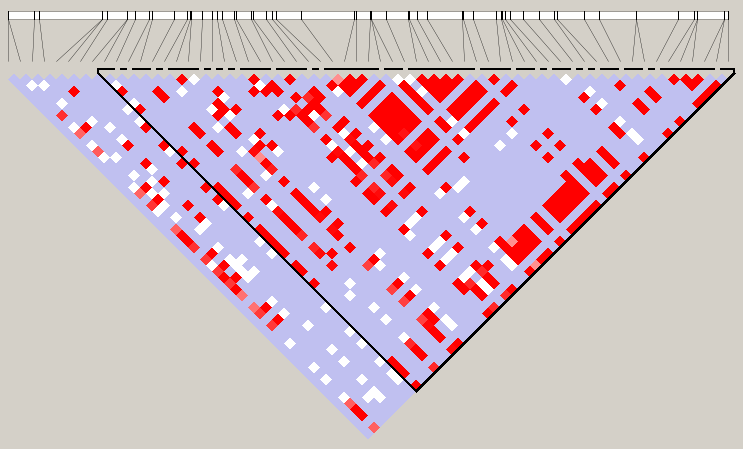 |
| 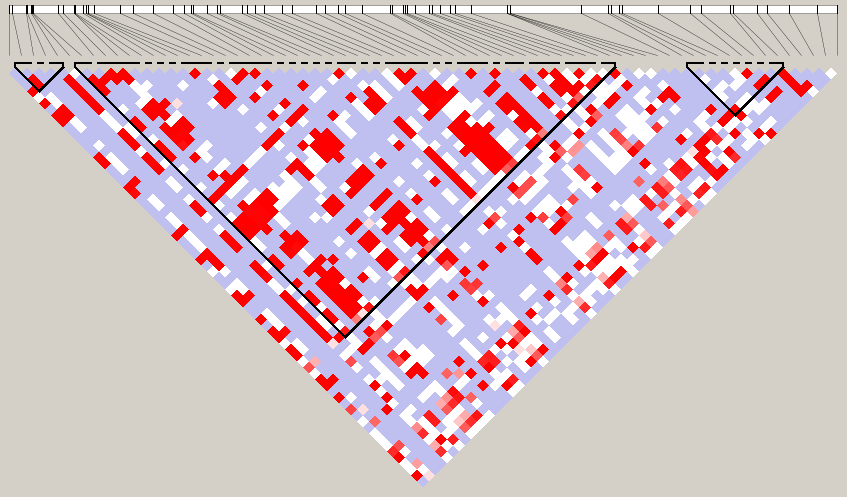 | 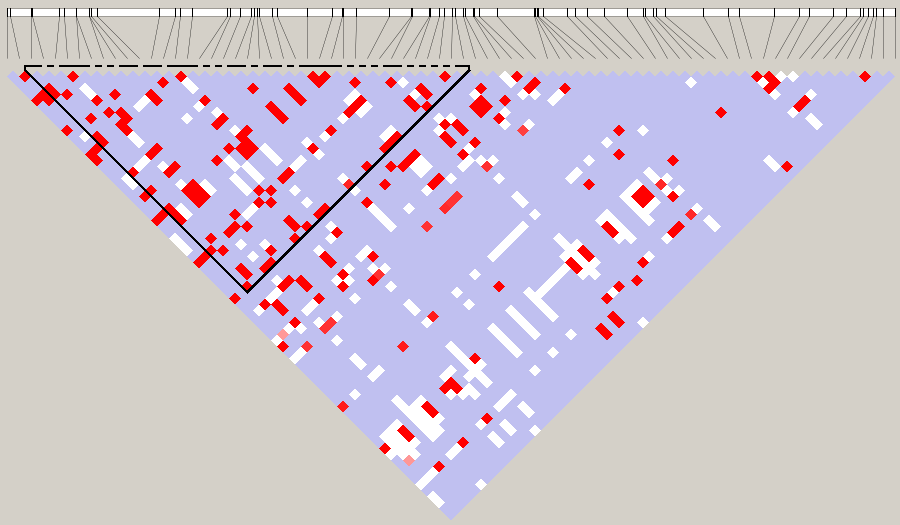 |
| A2 |  |
| 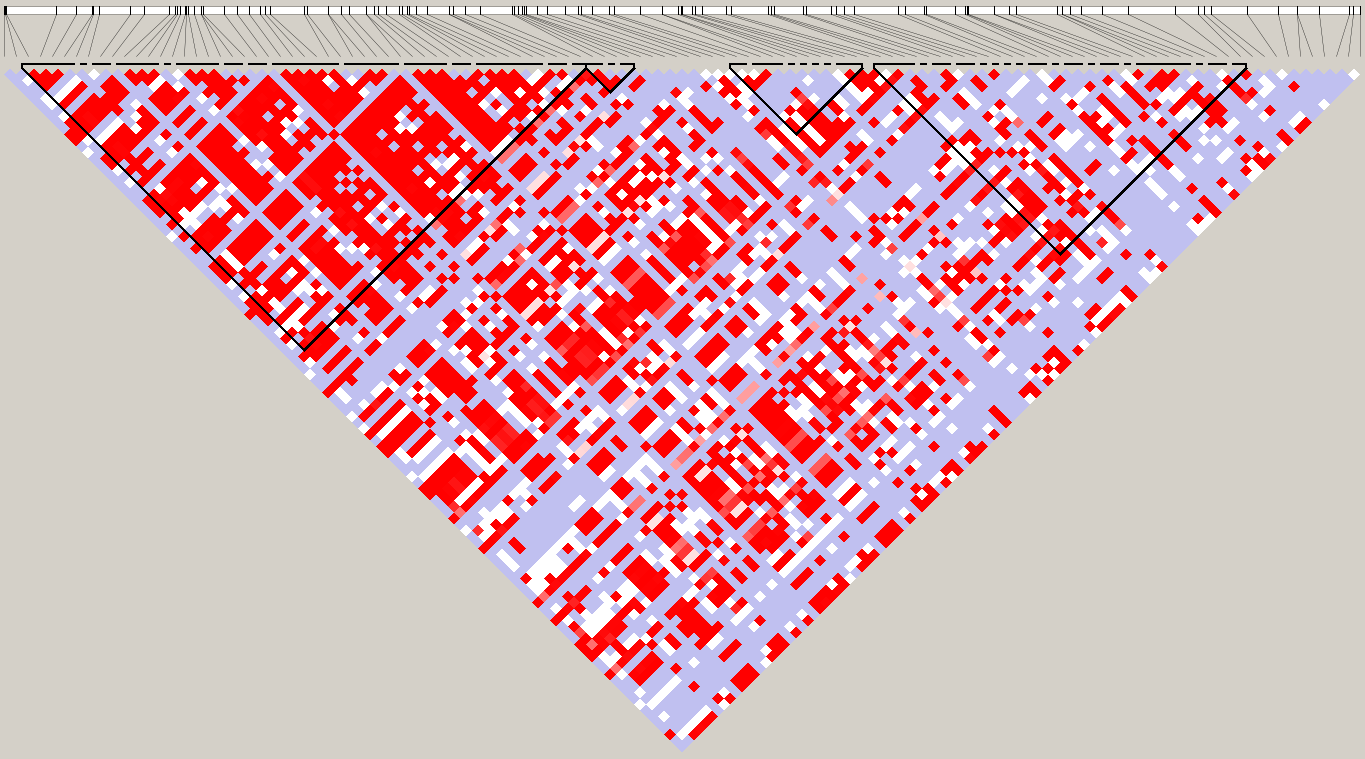 | 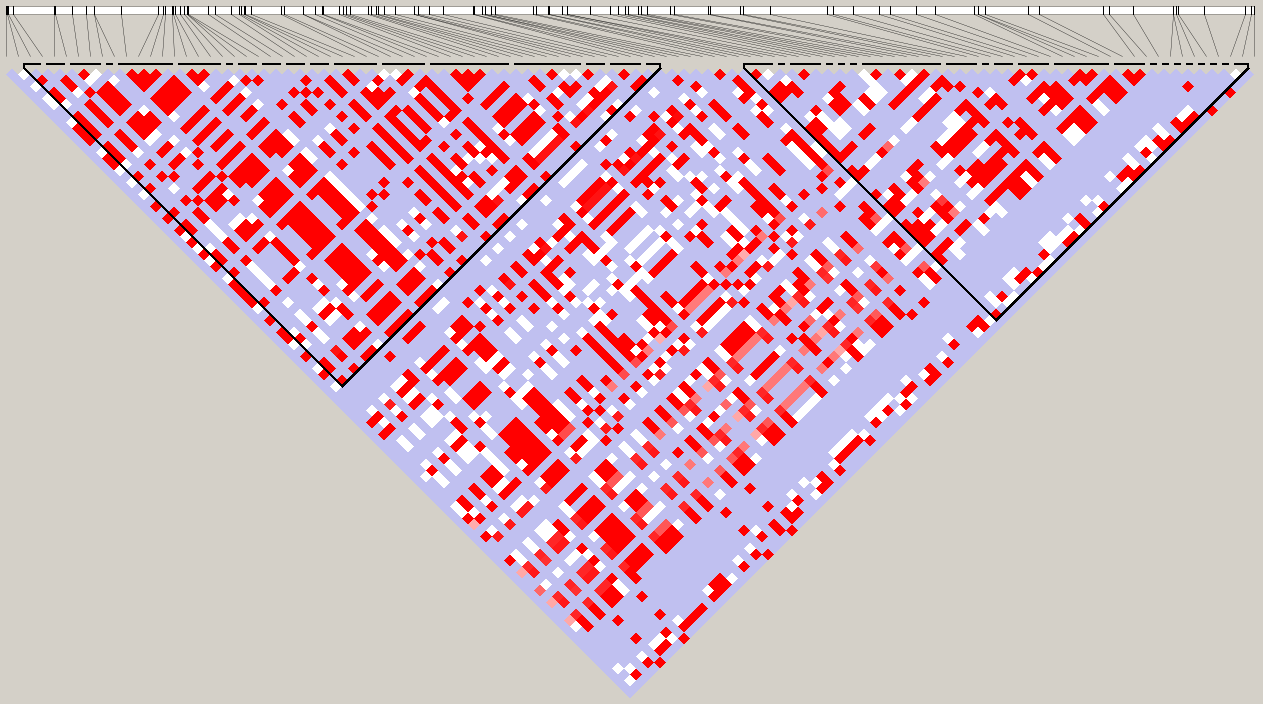 |
| A4 |  |
| 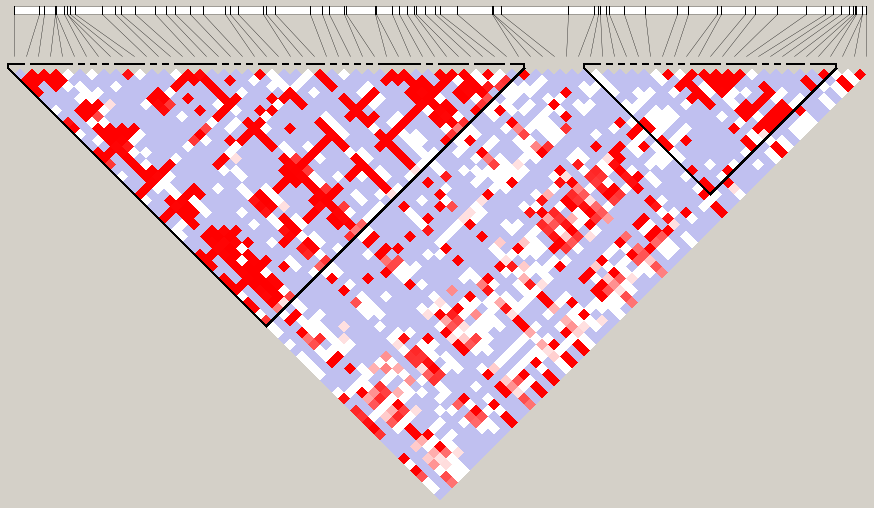 | 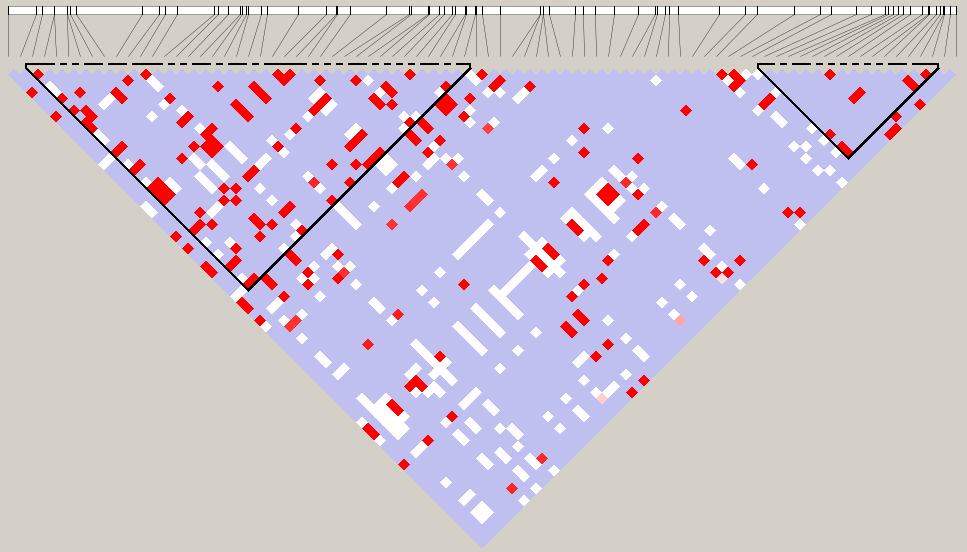 |
| B1 |  |
| 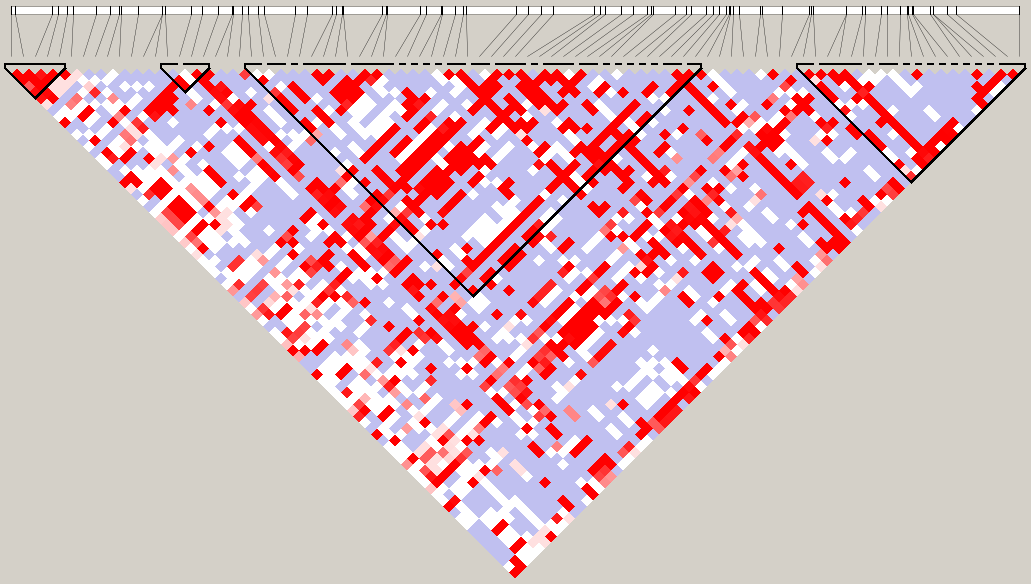 | 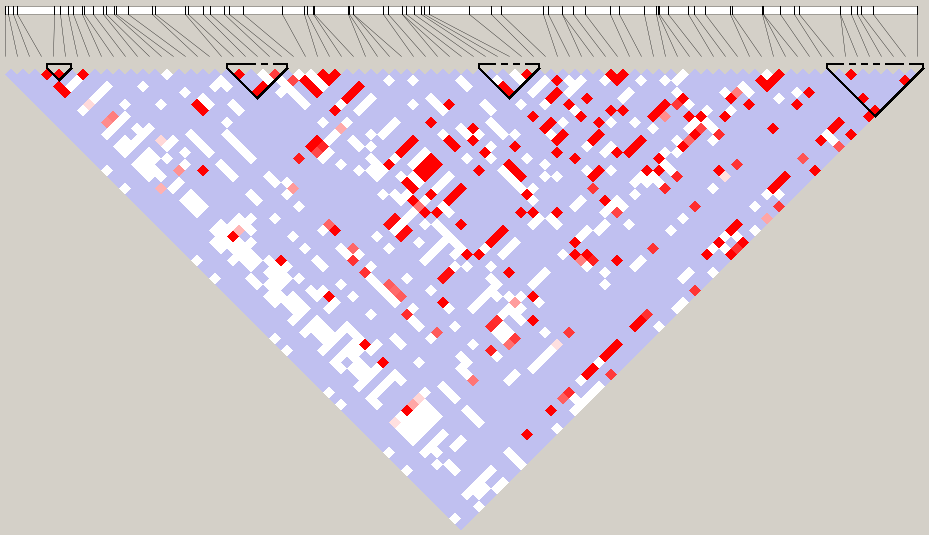 |
| B2 |  |
| 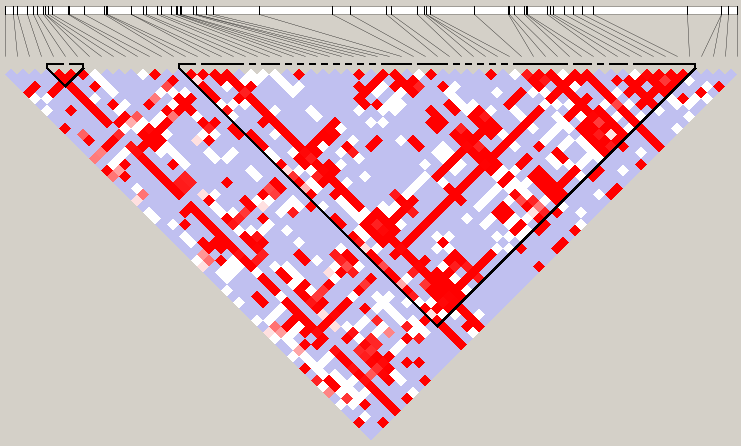 | 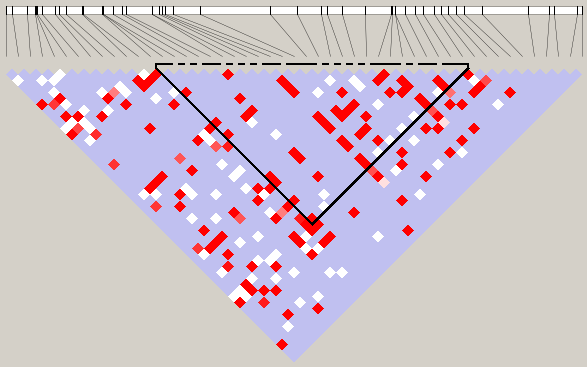 |
| B2 |  |
| 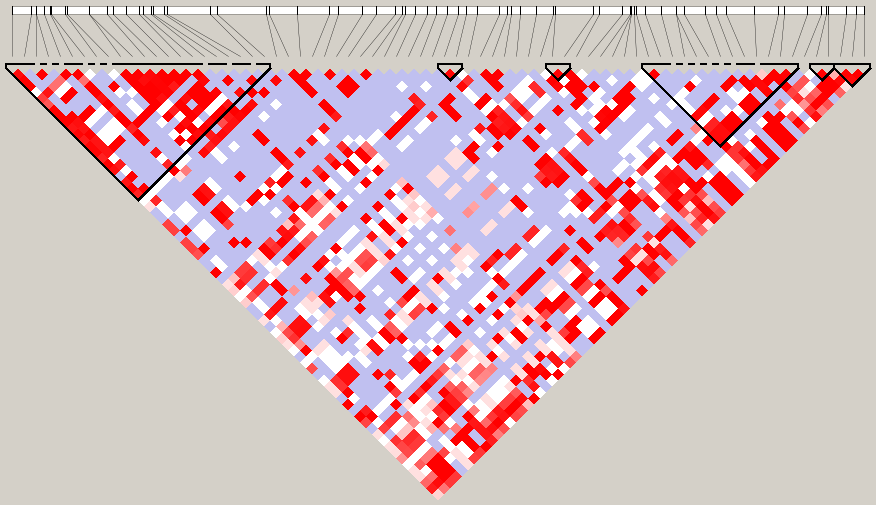 | 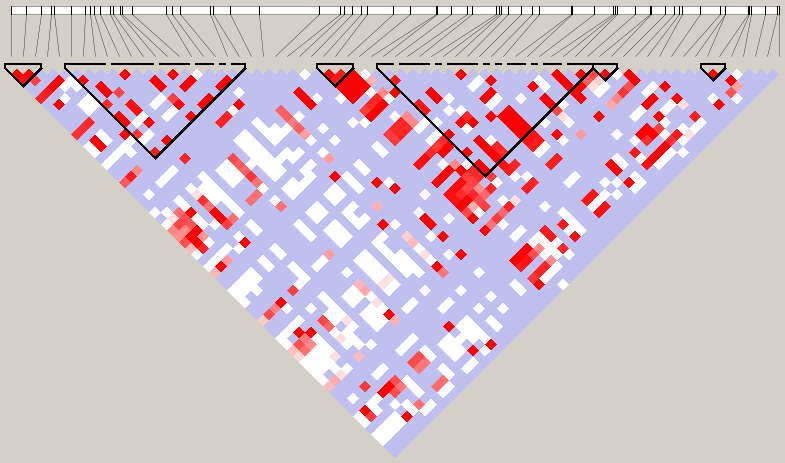 |
